# Discovery and validation of non-canonical antigens for Hepatocellular Carcinoma immunotherapy

**DOI:** 10.1101/2025.06.08.25328714

**Authors:** Stephen Li, Yujuan Dong, Jiaxun Liu, Shanglin Li, Dandan Pu, Yue Guo, Chun Kwok Wong, Nathalie Wong, Jason Wing Hon Wong

## Abstract

Hepatocellular carcinoma (HCC) is a highly prevalent and lethal form of liver cancer marked by high rates of tumor recurrence, posing substantial challenges to primary treatment options such as surgical resection and liver transplantation. Immunotherapy, which leverages T-cell-mediated recognition of tumor-specific antigens, has emerged as a promising alternative therapeutic strategy. While proteogenomics and ribosome profiling data across cancer types have provided growing evidence of active translation in non-canonical open reading frames (ncORFs), antigens derived from HCC-associated ncORFs remain largely unexplored. By integrating long-read and short-read RNA sequencing data with mass spectrometry, we have successfully identified twelve non-canonical peptides arising from cryptic translation of ncORFs. ELISpot assays confirmed cytokine release triggered by non-canonical peptides originating from distinct genomic loci. Subsequent RNA level analysis revealed four peptides with strong evidence of enriched expression in tumorigenesis. In-depth bioinformatics predictions indicate that these four peptides possess significant binding affinity for Major Histocompatibility Complex and T-Cell Receptor, with p2 emerging as the most promising candidate. Analysis using public datasets and experimental validation further highlighted the role of ribosome subunit recruitment and N6-Methyladenosine (m^6^A) modifications in enhancing translation initiation for tumor-enriched antigens. By addressing the existing shortage of knowledge in the HCC ncORF translation, this study identifies promising antigenic candidates for HCC immunotherapy, offering new avenues for effective treatment strategies and improved patient outcomes.

## Introduction

Hepatocellular carcinoma (HCC) is a highly prevalent form of primary liver cancer and a leading cause of cancer-related mortality [1]. The development of HCC is closely linked to several risk factors, including chronic infections of Hepatitis B virus (HBV) and Hepatitis C virus (HCV), alcohol abuse, and metabolic dysfunctions such as non-alcoholic fatty liver disease (NAFLD) [2, 3, 4, 5]. Surgical resection remains the primary treatment for early-stage HCC, though eligibility is dependent on factors such as adequate liver function, tumor size, and absence of extrahepatic metastasis [6]. Nonetheless, 70% of HCC surgery patients experience tumor relapse within five years [7]. Targeted therapies utilizing multi-target kinase inhibitors have emerged as an alternative for patients with unresectable HCC. The US Food and Drug Administration (FDA) has approved sorafenib and lenvatinib as first-line targeted treatments based on prolonged patient survival in clinical trials [8, 9, 10, 11]. Second-line therapies such as regorafenib, cabozantinib, and ramucirumab have also received FDA approval for patients previously treated with sorafenib [12, 13, 14]. Despite the advancements, these treatments provide only modest clinical benefits, underscoring the urgent need for more effective therapeutic strategies.

In recent years, immunotherapy has demonstrated significant potential in reshaping the HCC therapeutic landscape. The current standard first-line intervention for advanced HCC integrates a PD-L1 inhibitor atezolizumab with the anti-vascular endothelial growth factor monoclonal antibody (mAb) bevacizumab. The landmark IMbrave150 phase III trial in 2020 revealed that combination therapy with atezolizumab and bevacizumab offered clinical benefits superior to sorafenib, resulting in rapid FDA approval [15]. In 2022, the FDA approved another dual immune checkpoint blockade (ICB) HCC therapy of anti-PD-L1 mAb durvalumab with the anti-CTLA-4 mAb tremelimumab, based on the improved median overall survival and objective response rates over the sorafenib treatment in the phase III HIMALAYA trial [16].

The effectiveness of anti-tumor immunity largely depends on T-cell recognition of MHC I-associated peptides (MAPs). Ribo-Seq and proteomic evidence have suggested that non-coding regions, such as frame-shift ORFs, long non-coding RNAs (lncRNAs), and untranslated regions (UTRs), constitute an underexplored reservoir of MAPs [17, 18, 19, 20]. However, despite their potential significance, no immunopeptidome dataset for HCC is available in the pan-cancer non-canonical MHC I-associated peptides (ncMAPs) database IEAtlas [19]. Therefore, our objective is to characterize HCC ncMAPs through the integrative analysis of transcriptomic and proteomic data derived from our in-house established HCC cell models.

In a previous study, we combined PacBio SMRT long-read sequencing and Illumina short-read RNA-Sequencing to identify tumor-specific transcripts using in-house HCC cell models (HKCI) established from patients’ tumorous liver tissue [21, 22, 23]. Due to the higher tumor purity of HKCI cell models compared to primary HCC tissue samples, HKCI cell lines provide a strategic advantage in detecting low abundance ncMAPs. In this study, we further investigated the HCC immunopeptidome of the HKCI models using MHC-pulldown mass spectrometry [24]. By searching the HKCI-specific transcripts against the mass spectrometry data, we identified twelve distinct ncMAPs. We then employed computational and experimental approaches to select ncMAPs that exhibit the highest potential as neoantigen (**Figure 1**A). Our results have generated valuable datasets for understanding the HCC ncMAPs landscape and identified promising neoantigen candidates for future HCC immunotherapy development.

**Figure 1:**
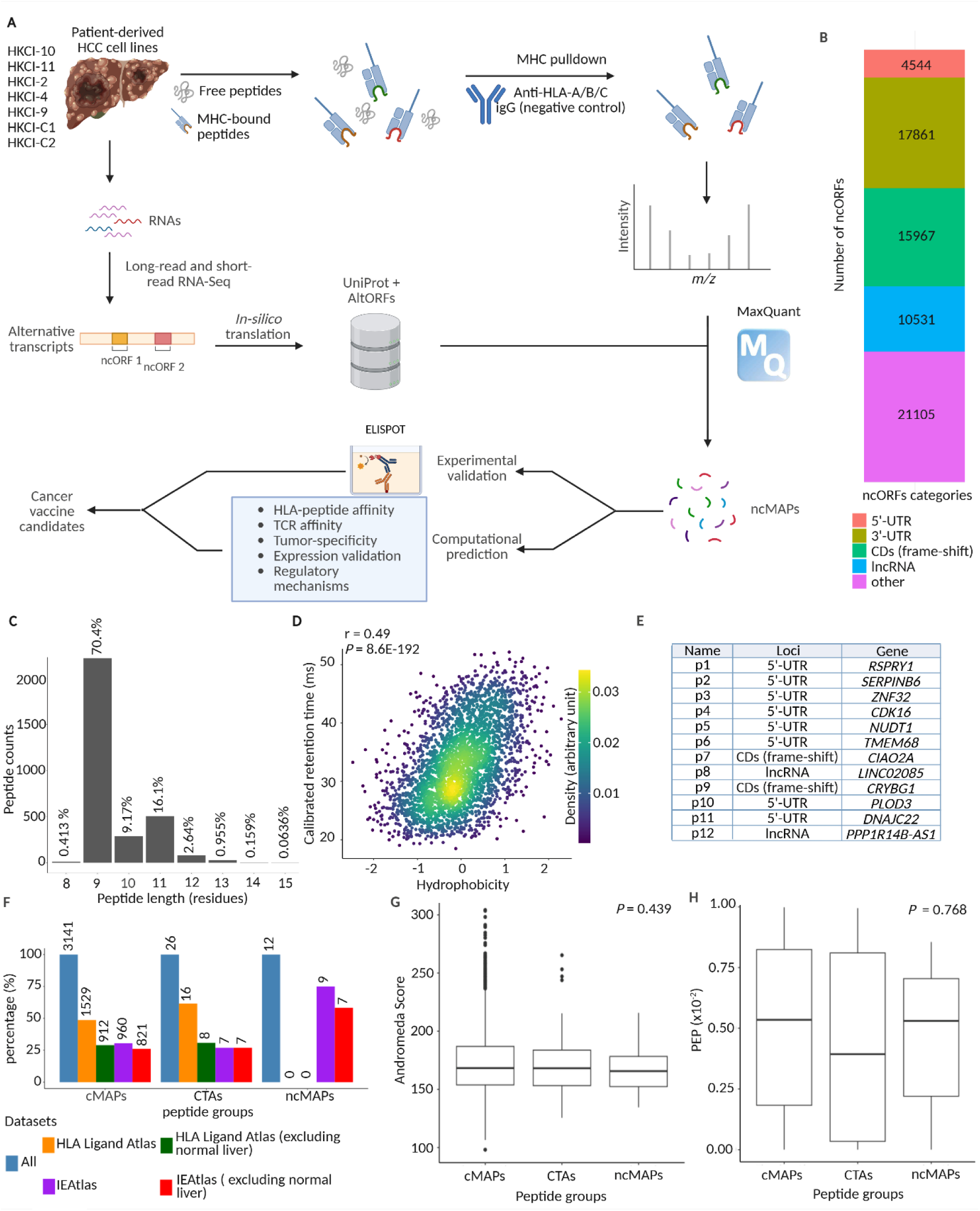
Overview of immunopeptidome data generated from HCC patients. **A.** Workflow of the proteogenomic search of ncMAPs using ncORFs, and subsequent downstream selection of candidates for development of cancer vaccine. **B.** ncORFs in the customised proteomics databases originated from various genomics regions. **C.** Distribution of peptide lengths observed in the generated immunopeptidome. **D.** Correlation between calibrated retention time and predicted hydrophobicity. **E.** Names, genomic origins and associated genes of the twelve identified ncMAPs. **F.** Proportion of identified ncMAPs found in database searches against HLA Ligand Atlas and IEAtlas, and their presence in normal liver samples. The text labels above each bar indicate the number of identified peptides. **G.** to **H.** Distribution of MaxQuant Andromeda Score and PEP across all cMAPs, CTAs, and ncMAPs. ncMAPs, non-canonical MHC-associated peptides; ncORFs, non-canonical open reading frames; PEP, posterior error probability; CTAs, cancer-testis antigens.

## Results

### Mass spectrometry identifies canonical and non-canonical MHC-I associated peptides

To facilitate the identification of ncMAPs, we employed anti-MHC affinity purification to selectively enrich ncMAP-associated peptides, followed by mass spectrometry analysis (Figure S1A). Cancer cells frequently downregulate MHC class I molecules to facilitate immune evasion. Before conducting MHC-based affinity purification, we used flow cytometry to evaluate the surface HLA-A/B/C expression in HKCI cell lines. Analysis of the mean fluorescence intensity showed a significant signal in all HCC cell lines stained with HLA-A/B/C antibody (clone W6/32) compared with isotype control, confirming that all seven cell lines were MHC class I-positive, thereby validating their suitability for systematic tumor antigen profiling (Figure S1B). In addition, three stringent quality control measures ensured sufficient IgG/antibody loading, optimal binding of antigen-HLA complexes, and effective antigen elution from the column (Figure S1C, D and E)

The identification of canonical and non-canonical MAPs from the seven HKCI cell lines was achieved by integrating transcriptomic and proteomics data (**Figure 1**A). To clarify the source of the ncORFs, we extracted ncORF RNA sequences and aligned them using Minimap2. The resulting alignment files were annotated with hg19.ncbiRefSeq.gtf to classify the genomic origin of individual ncORFs. ncORFs located in unannotated regions or overlapping with start or stop codons were categorized as “others”. The raw mass spectrometry data was analyzed against protein databases that included the UniProt reference and 70,008 ncORF sequences identified in HCC cell lines. Among the 70,008 detected ncORF sequences, 21,105 were located in unannotated regions, while 5’-UTR ncORFs represented the smallest subset, accounting for only 4,544 sequences (**Figure 1**B). Integrating UniProt reference sequences with customized databases improves the accuracy of novel peptide identification by reducing false positives. This approach mitigates errors arising from identical or similar sequences within UniProt, as well as distinct sequences modified post-translationally, thereby enhancing overall reliability.

The peptide filtering step described in the Materials and methods section was implemented to ensure the statistical robustness, MS/MS spectra validation, and biological relevance of the remaining candidates. 70.4 % of peptides measured nine amino acids in length, consistent with HLA-I enriched peptides (**Figure 1**C). Moreover, predicted hydrophobicity exhibited a positive correlation with retention time in the liquid chromatography column (**Figure 1**D), further substantiating the peptide identification by MaxQuant. After filtering out peptides that fell outside the expected residue length range for MHC-I-associated peptides or failed statistical validation, a total of 3,141 peptides were retained. Among them were 26 Cancer-Testis Antigen (CTA) peptides that are tumor-associated antigens exclusively expressed in malignant tissues and normal testis. Notably, the mass spectrometry data also revealed twelve novel ncMAPs (p1 – p12) (**Figure 1**E, Table S1). The theoretical MS/MS spectra of the ncMAPs closely aligned with the experimental spectra, reinforcing the reliability of their identification (Figure S2).

To evaluate whether the peptides detected in this study are present in previous mass spectrometry data, and mitigate misidentification arising from redundant translation of canonical reference peptides as non-canonical, we compared all identified peptides against public immunopeptidome databases, including HLA Ligand Atlas which covers the UniProt proteome, and IEAtlas which focuses on peptides derived from ncORFs. Approximately half of the cMAPs (1,529 out of 3,141) were cataloged in the HLA Ligand Atlas. The absence of the remaining peptides may indicate either the lack of detectable MHC presentation or removal by the HLA Ligand Atlas’ pre-filtered step based on theoretical HLA affinity. Nevertheless, canonical MAPs (cMAPs), including CTA, were more frequently found in the HLA Ligand Atlas than in IEAtlas. cMAPs identified in IEAtlas may stem from codon degeneracy, producing peptide sequences indistinguishable from ncORF-derived products. In contrast, nine out of twelve identified ncMAPs were present in IEAtlas, supporting the existence of these candidates. Furthermore, the absence of ncMAPs in the HLA Ligand Atlas confirmed that all twelve candidates are authentic ncMAPs, rather than artefacts of degenerated codon translation from canonical ORFs. (**Figure 1**F).

We then focused on the immunopeptidomics data specifically in normal liver tissues to assess the risk of non-tumor antigen recognition. 912 out of 1,529 cMAPs detected in the HLA Ligand Atlas were absent in normal liver. We then assessed the presence of ncMAPs in adjacent normal tissues using publicly available immunopeptidomics data compiled in IEAtlas. Seven ncMAPs from IEAtlas were absent in normal liver tissues in IEAtlas, indicating their potential as antigen candidates with minimal risk of triggering host immunity in the liver (**Figure 1**F). Furthermore, the novel ncMAPs exhibited Andromeda scores - a metric that quantifies the quality of the peptide spectrum (PSM) - that were statistically comparable to those of all cMAPs and CTAs (**Figure 1**G). Moreover, ncMAPs showed no significant deviation from cMAPs and CTAs in posterior error probabilities (PEP), which quantify the statistical likelihood of false PSM identification (**Figure 1**H). The consistency of these metrics between ncMAPs and cMAPs/CTAs reinforces the robustness of mass spectrometry-based identification.

### Candidate ncMAPs from diverse genomic origins induce CD8^+^ T cell activation

Each HKCI cell line expressed and presented at least three ncMAPs. While some ncMAPs were only present in one sample, other ncMAPs — such as p12 — underwent recurrent translation across multiple samples (**Figure 2**A). The peptide length distribution closely mirrors that of the overall immunopeptidome, with over half of the ncMAPs being nine amino acids long (**Figure 2**B). Interestingly, despite being the least abundant category in the customized ncORF database, seven out of twelve candidate peptides were translated from 5’-UTRs (untranslated regions) (**Figure 2**B, **2**C and **1**B). For example, p2 is derived from the extended 5’ end of a long isoform of Serpin family B Member 6 (*SERPINB6*) (**Figure 2**D). Additionally, Transposable element (TE)-exon splicing provides an alternative mechanism for 5’-UTR translation, as exemplified by p10, which originates from the 5’-UTR of Procollagen-Lysine, 2-Oxoglutarate 5-Dioxygenase 3 (*PLOD3*). This 5’-UTR is initiated by Mammalian-wide Interspersed Repeat element 3 (*MIR3*), a member of Short Interspersed Nuclear Element (SINE) (**Figure 2**E). Similarly, p5 arises from the 5’-UTR of Nudix Hydrolase 1 (*NUDT1*), annotated within the long-terminal repeat region (*ERV1*) (**Figure 2**F). Notably, the p5 ORF spans splice junctions between two exons of *NUDT1*, underscoring splice junctions as a potential source of ncORF translation. Likewise, p3 arises from the 5’-UTR of Zinc Finger Protein 32 (*ZNF32*), further illustrating splice-junction-derived ncORFs (**Figure 2**G). In addition to the 5’-UTR, p8 and p12 originate from translation events of lncRNAs (Figures 2C and 1E). For instance, p8 is derived from the immunoregulatory lncRNA *LINC02085*, indicating a dual immunoregulatory function at both the RNA and peptide levels (**Figure 2**H). Moreover, three peptides originated as frame-shifted translation products from protein-coding exons. For instance, p9 is a frame-shifted CDS-derived peptide, generated from Crystallin Beta-Gamma Domain Containing 1 (*CRYBG1*), a putative tumor suppressor in melanoma [25] (**Figure 2**I).

**Figure 2:**
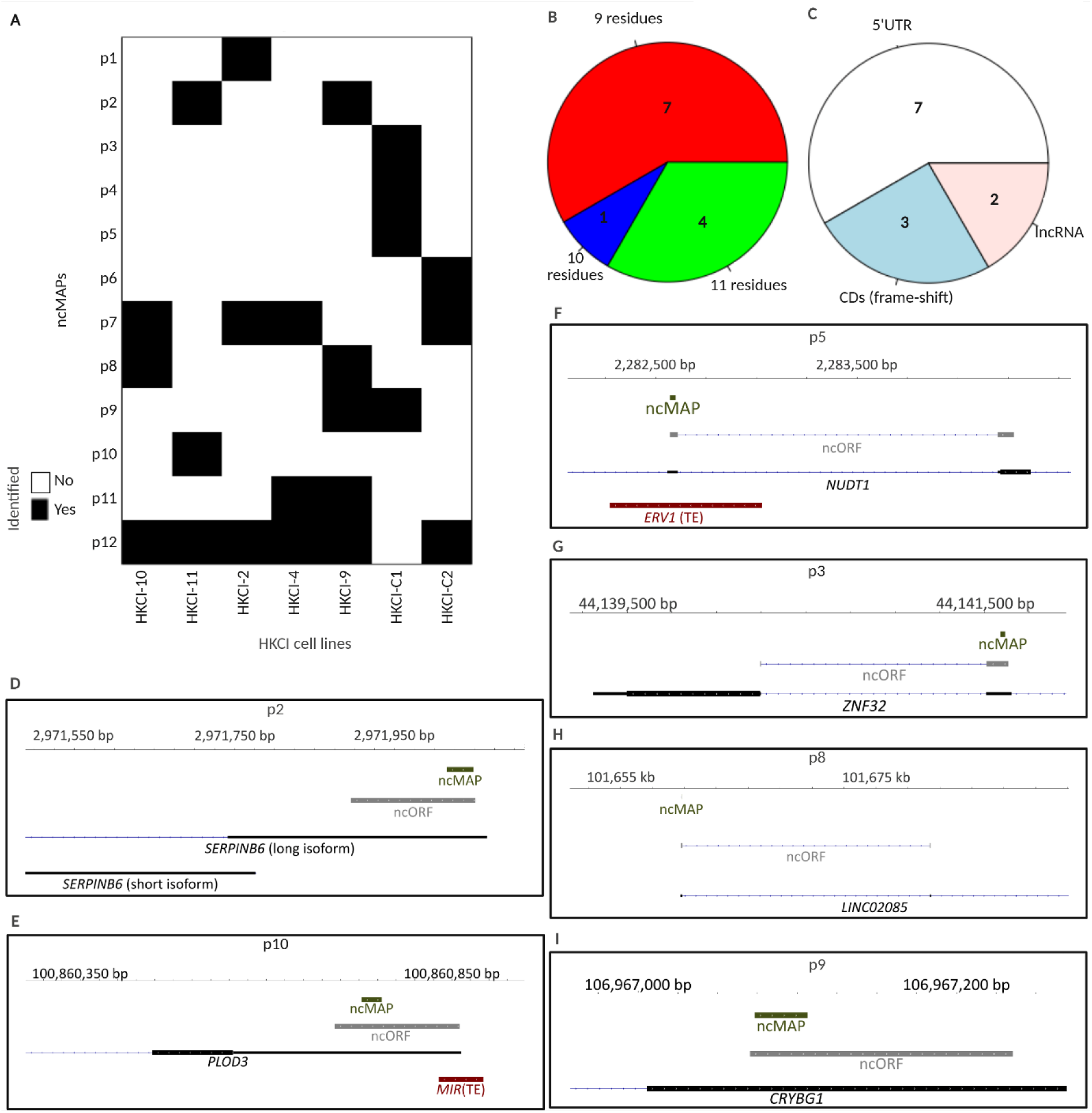
Expression and visualization of the twelve novel ncMAPs. **A.** Distribution of the twelve ncMAPs candidate peptides identified across the HKCI cell lines **B.** Distribution of read lengths for the ncMAPs candidate peptides. **C.** Classification of the ncMAPs candidate peptides based on their genomic positions. **D.** to **G.** IGV visualization of the representative ncMAPs and ncORFs derived from the 5’-UTR. **H.** IGV images displaying lncRNA-derived ncMAP p8 and its ncORFs. **I.** IGV image displaying ncMAP p9 produced from a frameshift ncORF of a canonical exon. IGV, Integrative Genomics Viewer; lncRNAs, long non-coding RNAs; ORFs, open reading frames; TE, transposable element.

To emphasize the importance of PacBio long-read sequencing in identifying splice variants, we examined transcripts associated with p3 and p5 to visualize read alignment. The ORFs of p3 and p5 span splice junctions, illustrating the unique advantage of long-read sequencing in precisely resolving these junctions and their corresponding ncORFs. The circular consensus sequencing (CCS) data from HKCI-4 and HKCI-C1 validated the *ZNF32* transcripts, which include the splice junction covered by the p3 ORF (Figure S3A). Similarly, HKCI-4 yielded a CCS read validating the *NUDT1* isoform linked to the p5 ORF (Figure S3B). These findings underscore the critical role of long-read sequencing in reliably identifying isoforms. Collectively, our findings emphasize the diverse origins and molecular features of the peptides analyzed in this study.

To verify that the ncMAPs identified in our cell line model are also translated and presented in heterogeneous primary HCC tissue samples, we re-analyzed the MHC-I peptidome (PXD023143) and observed that peptides p1, p8, and p9 were also presented in the tumor tissues of HCC patients (Figure S4A). Furthermore, we examined the presence of the twelve ncMAPs in a second immunopeptidomics dataset from HBV-positive HCC patients (PXD037270) and identified the presence of nine ncMAPs in a cohort of 48 HCC patients (Figure S4B).

The twelve ncMAPs exhibit varying degrees of novelty based on their genomic origins, which may influence their potential as neoantigens. Peptides arising from frame-shift mutations or splice junction derived ncORFs may generate more novel sequences than those originating from UTRs. We utilized the ELISpot assay to quantify cytokine Interferon gamma (IFN-*γ*) released by CD8^+^ T cells that were co-cultured with antigen-presenting cells, which were pulsed with candidate peptides encoded from various genomic loci. We selected three peptides for immunogenicity analysis: p2, an example of ncMAPs derived from 5’-UTR; p9, representing a frame-shift product; and p3, selected as a representative of junction-spanning ORFs (**Figure 3**A). ELISpot analysis was conducted using peripheral blood mononuclear cells (PBMC) isolated from six healthy donors. Our ELISpot result showed that CD8^+^ T cell responses to the cancer-testis antigen NY-ESO-1 (positive control) but not the Actin_1-9_ peptide (negative control) at a concentration of 1 µg/mL. This pattern confirms the more pronounced immune activation signals induced by MHC-I presented antigens over background controls (**Figure 3**B and C). Our findings revealed that the three candidate peptides induced significantly stronger T cell activation over Actin_1-9_ in at least three of six donors. Among the candidate peptides, p9 consistently induced IFN-*γ* secretion in all six donors. Compared to p9, p2 and p3 triggered significant amount of IFN-*γ* production in donors 5 and 6, and surpassed effect of NY-ESO-1 peptide under identical concentration and treatment conditions (p-value *<* 0.01). Overall, our results suggest that frame-shift products elicit a robust immune response, while peptides derived from extended 5’-UTRs and splice junction-associated ncORFs can also induce substantial cytokine release, supporting the immunogenic potential of the candidate ncMAPs.

**Figure 3:**
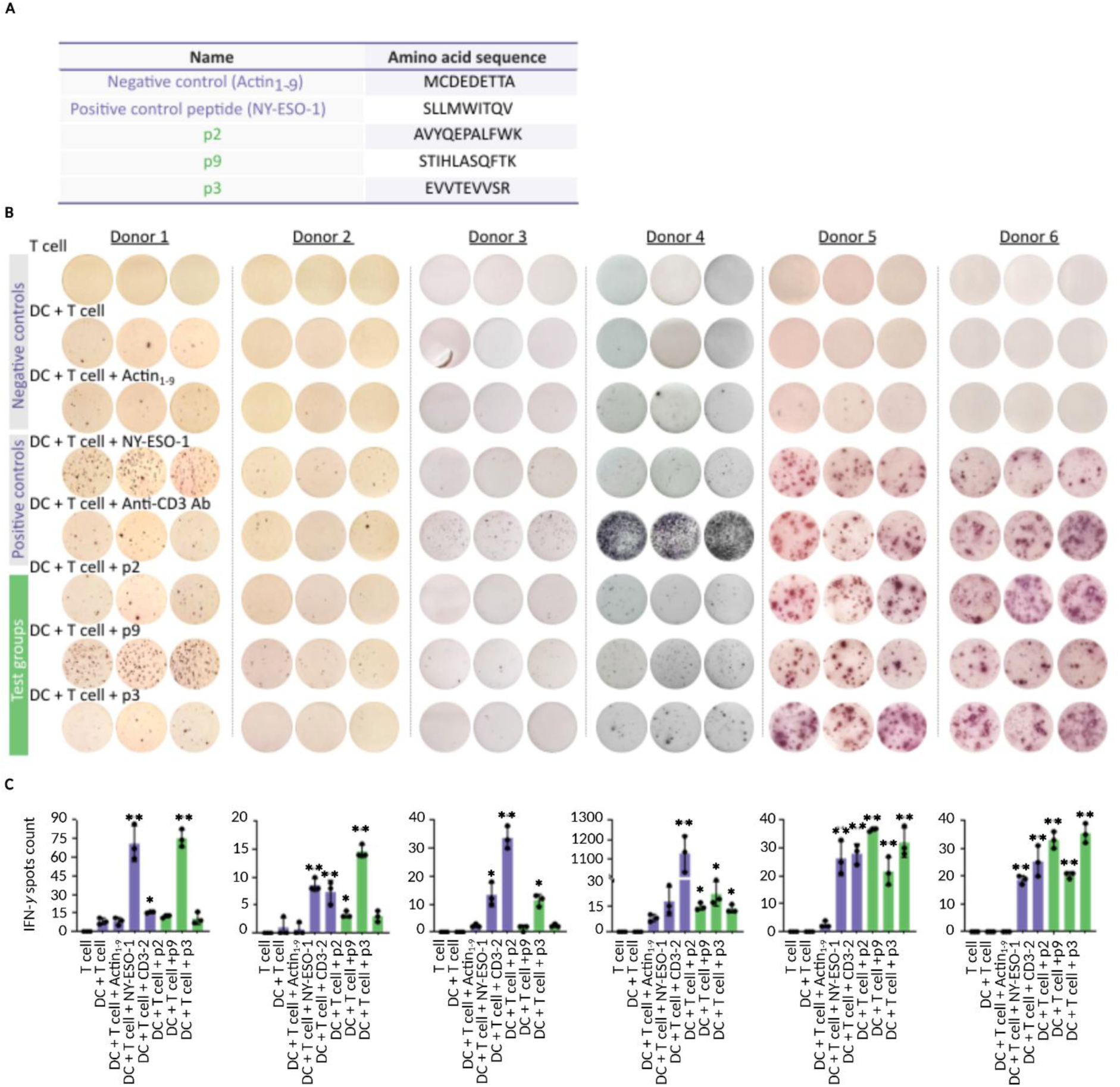
Validation of the immunogenicity of candidate peptides using IFN-*γ* ELISpot assay. **A.** Peptides tested in the ELISpot assay. **B.** Images of the ELISpot assay showed the IFN-*γ* secretion by DC:CD8^+^ T cells co-culture following 24 hours stimulation with various peptides. Autologous DC and CD8^+^ T cells isolated from healthy donors were used in the analysis. **C.** Summary of the IFN-*γ* spot number. Statistical analysis was performed against Actin_1-9_ peptide using unpaired t-test. Data is presented as mean values ± SD. \**p<*0.05, \*\**p<*0.01. ELISpot, enzyme-linked immunosorbent spot; DC, dendritic cell; IFN-*γ*, interferon gamma.

### RNA-level abundance identifies peptides with the strongest evidence of tumor enrichment

To prioritize candidates for development of potential immunotherapy, we assessed the twelve ncMAPs by examining their tumor specificity at the RNA expression level. We screened exon expression using GTEx dataset and found a low abundance of ncORF expression in normal liver and other normal tissue types. For example, the exon containing the p2 ORF exhibited restricted expression across the liver and other tissue types in GTEx (Figure S5A). Likewise, the exon harboring p9 showed a modest expression level across GTEx liver tissues and most other tissue types (Figure S5B).

For a more comprehensive assessment of the abundance ncORF transcript in clinical HCC specimen (T) and adjacent normal liver tissues (AN), we employed RT-qPCR quantification in a cohort of 32 HCC patients. Our analysis showed that approximate 50% HCC patients had at least five ncORFs upregulated in the tumor tissues, with a T/AN ratio exceeding 1.5-fold (**Figure 4**A). 87.5% of patients harbor at least one ncORF upregulated in the tumors (**Figure 4**A). Candidate peptides p2, p4, p5, p6, p10, and p12 exhibited frequent overexpression across patient samples (Figure 4A). Notably, p2, p4, p5, p6, and p10 showed statistically significant tumor upregulation (**Figure 4**B).

**Figure 4:**
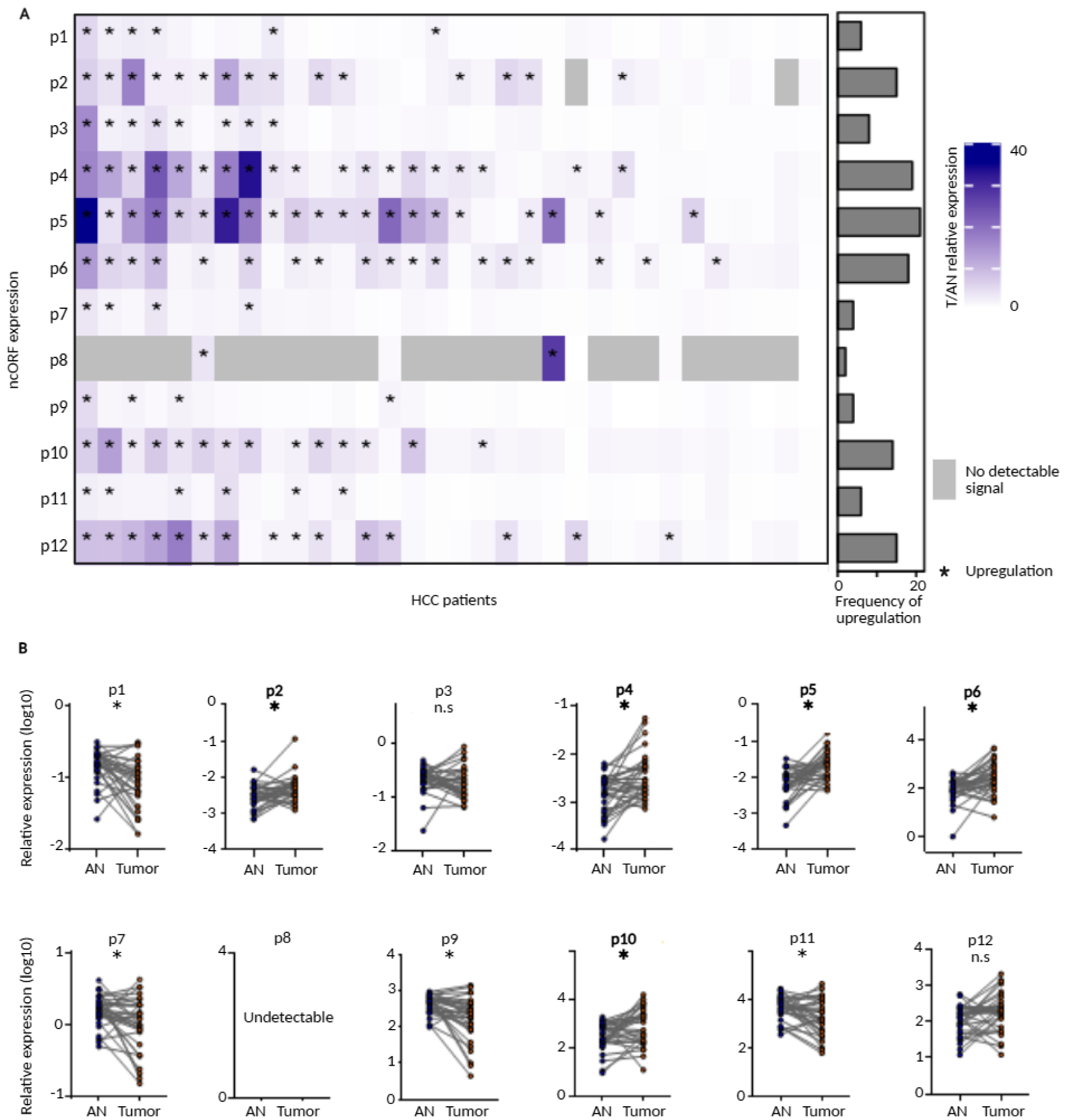
Detection of ncORF expression in clinical HCC specimens using RT-qPCR. **A.** Ratio of relative ncORFs expression between clinical HCC tumor and matched adjacent normal liver tissues across 32 HCC patients, ordered by the frequency of upregulation (T/AN RT-qPCR relative expression fold-change *>* 1.5. **B.** Statistical analysis of the expression of twelve ncORFs. Data was analyzed using paired T-test. \**p<*0.05, \*\**p<*0.01. *n.s.*, not significant. T/AN, Tumor to normal adjacent tissue; RT-qPCR, Quantitative reverse transcription polymerase chain reaction; HCC, Hepatocellular carcinoma.

RT-qPCR is prone to errors due to primer binding to only a portion of the ncORF, which may result in amplification bias and reduced efficiency. Moreover, the limited sample size of our current HCC cohort could affect the statistical power and reliability of conclusions. To mitigate these constraints, we complemented RT-qPCR analysis of 12 ncORFs with RNA-seq dataset of GTEx liver and TCGA-LIHC samples, allowing for a comparative assessment across a larger cohort with full ncORF transcript coverage. Except for p8, most ncORFs exhibited read coverage across the majority of samples, with p5 and p6 displaying the most pronounced upregulation in TCGA-LIHC (**Figure 5**A). This observation was further confirmed with statistical comparisons, revealing several additional candidates associated with significant upregulation in HCC tumors, including p1, p2, p3, p5, p6, p7, and p10 (**Figure 5**B). Collectively, our analysis provided solid evidence supporting the HCC tumor enrichment of candidates p2, p5, p6, and p10. These findings justify further experimental evaluation of their immunogenic potential.

**Figure 5:**
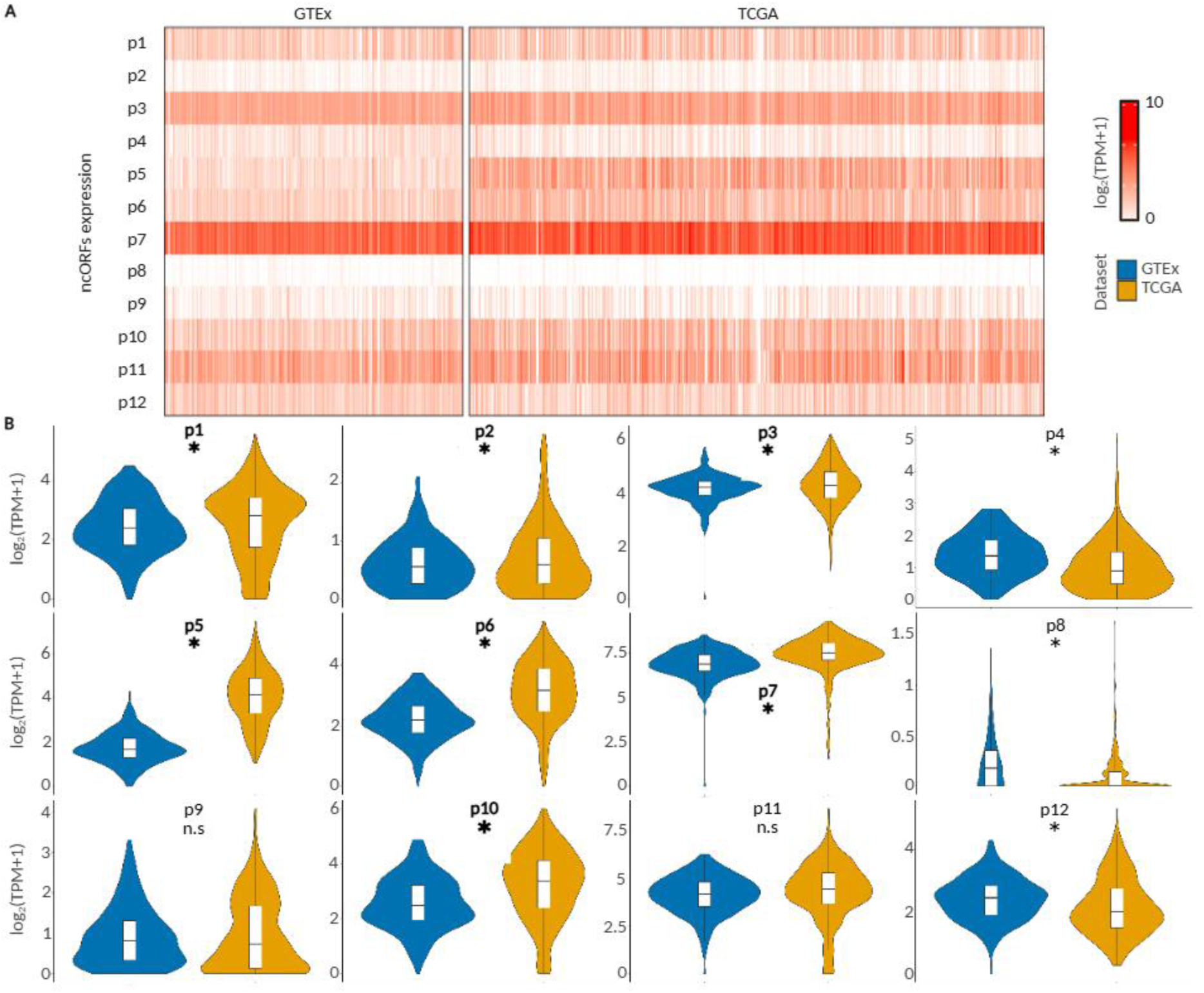
Expression of ncORFs in publicly available transcriptomic datasets. **A.** Log-transformed TPM expression of the twelve ncORFs in GTEx liver and TCGA-LIHC samples. **B.** Grouped statistical analysis of the RNA-Seq expression of the twelve ncORFs.. \**p<*0.05, \*\**p<*0.01. TPM, transcript per million; TCGA, The Cancer Genome Atlas; LIHC, Liver Hepatocellular Carcinoma; GTEx, Genotype-Tissue Expression.

### Computational predictions reveal recurrent affinities to HLA across patients

To evaluate the HLA binding affinity of ncMAPs, we conducted HLA-typing for HKCI cell lines using PCR based method. Due to the limited resolution of PCR-based HLA typing, multiple HLA alleles may be assigned to the same cell lines. For instance, HKCI-9 was assigned HLA-A11:01, HLA-A11:02, and HLA-A11:04 due to the high similarity of the alleles DNA sequences indistinguishable by PCR typing. Similarly, HLA-C03:02, HLA-C03:04, and HLA-C03:06 were assigned to HKCI-C1. Moreover, multiple HLA allele subtypes with highly similar sequence and antigen-binding properties are sometimes classified under the same HLA supertype. In this study, several HLA-A02 subtypes (e.g., HLA-A02:01, HLA-A02:02, HLA-A02:04) were all assigned into the HLA-A02 supertype. Nonetheless, assigning multiple similar HLA alleles to a single cell line has minimal to no impact on downstream NetMHCpan analysis, as their sequence similarities result in nearly identical antigen-binding preferences.

We clustered the HKCI cell lines based on their HLA allele subtype distributions to visualize similarities and distinctions in their profiles. Certain cell lines, such as HKCI-2 and HKCI-4, shared similar alleles sets. Indeed, HLA-A02 is one of the most frequent serological types that presented in Chinese population (>10%). Therefore 6 out of 7 HKCI cells were classified as A02 supertype. In contrast, HKCI-C1 displayed a distinct HLA repertoire compared to the other six cell lines and was the only one lacking the A02 supertype, likely due to its different ethnic origin. Peptide-HLA affinities were predicted using NetMHCpan, with cell-line specific MAPs queried against all HLA-alleles within the respective cell line (Figure S6A). The results showed that most peptides exhibited affinity for at least one host HLA allele, with strong binders comprising the most substantial fraction (**Figure 6**B and C). These findings affirm the immunogenic potential of the peptides analyzed in this study.

**Figure 6:**
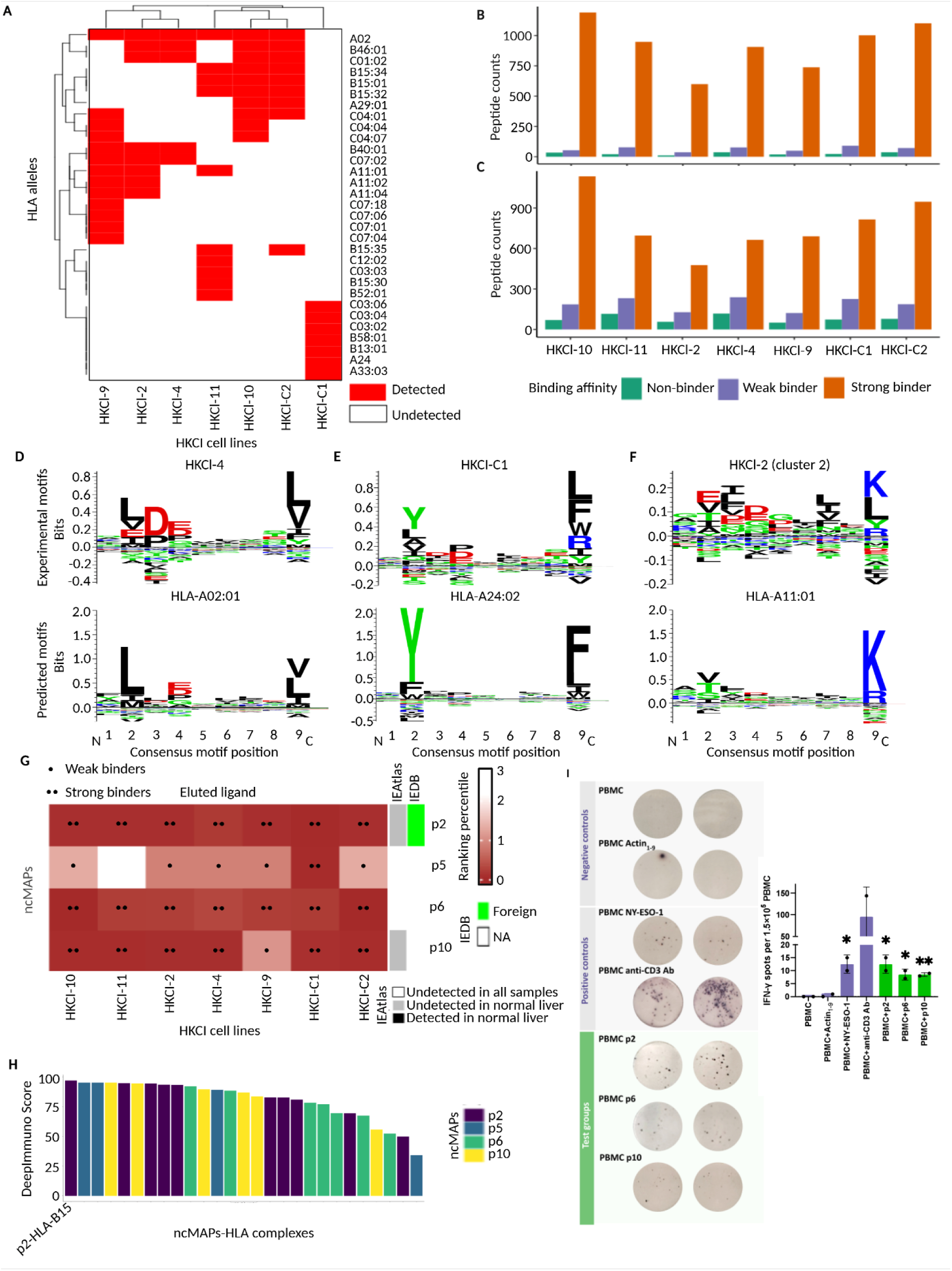
Immunogenic potential of identified MAPs. **A.** HLA serological types of HKCI cell models determined using LinkSeq HLA typing-kit. **B.** and **C.** NetMHCpan prediction of all identified MAPs in each HKCI cell lines based on EL and BA training datasets. **D.** Experimental consensus motif of HKCI-4 versus predicted motif of HLA-A02:01. **E.** Experimental consensus motif of HKCI-C1 versus predicted motif of HLA-A24:02. **F.** Experimental consensus motif of HKCI-2 versus predicted motif of HLA-A11:01. **G.** Patient-specific peptide NetMHCpan predictions from EL training dataset. Results from the BA training dataset are not presented here, as they closely resemble those from the EL training dataset. **H.** DeepImmuno evaluation of peptide-HLA interactions. **I.** Detection of immunogenicity of p2, p6 and p10 using ELISpot assay and PBMC of HCC patient. Statistical analysis was performed against Actin_1-9_ peptide using unpaired t-test. Data is presented as mean values ± SD. \**p<*0.05, \*\**p<*0.01. ELISpot, enzyme-linked immunosorbent spot; EL, eluted ligand; BA, binding affinity.

The patient-specific immunopeptidome identified in this study was further compared to HLA allele binding motifs predicted by NetMHCpan. Overall, the consensus motifs are consistent with graphical representations provided by MHC Motif Viewer [26]. For example, the consensus motif from HKCI-4 exhibited a leucine in the second position and leucine/valine in the ninth position. These patterns align with the consensus motif of the HLA-A02:01 peptidome (**Figure 6**D). We observed similar patterns among other cell lines carrying the HLA-A02 allele, such as HKCI-C2 and HKCI-9 (Figure S6B and C). The consensus motif of HKCI-C1 was distinct from that of HKCI-4 and other cell lines due to its unique set of HLA alleles. The most notable feature of the HKCI-C1 consensus motif is the tyrosine and phenylalanine in the second and ninth positions, respectively. This pattern likely originates from HLA-A24, which displays identical enrichment of tyrosine and phenylalanine in these positions (**Figure 6**E). In the case of HKCI-2, apart from 422 peptides belonging to a motif similar to that of HKCI-4 (Figure S6D), an additional 158 peptides formed a distinct cluster, showing higher background noise. This cluster may arise from motifs unique to HKCI-2 and absent in HKCI-4, such as the HLA-A11:01 motif, which consists of a lysine residue in the ninth position (**Figure 6**F). Some cell lines, such as HKCI-10, displayed multiple consensus motif clusters (Figure S6E and F).

We focused on the cell-line-specific ncMAPs-HLA affinity, specifically analyzing p2, p5, p6, and p10, which showed tumor-enriched expression in both RT-qPCR and RNA-Seq datasets. Among these, p2, p6, and p10 were predicted to interact with alleles across all seven cell lines (**Figure 6**G). Notably, IEAtlas data further supported the absence of p2 and p10 in the immunopeptidome of non-tumor liver and other tissues (Figure 6G). The absence in the IEAtlas normal tissue immunopeptidomics data is insufficient to conclude the tumor specificity of ncMAPs, due to the low detection sensitivity of mass spectrometry based immunopeptidomics and the inherent technical limitations of antibody-based peptide enrichment. To further enhance tumor specificity analysis, we integrated IEAtlas immunopeptidomics data alongside RT-qPCR and RNA-Seq evidence. IEAtlas complements RNA-level quantification by providing critical evidence that target candidates are translated and presented on tumor MHC while remaining absent in non-tumor liver tissues.

IEAtlas also recorded homology between p2 and foreign immunogenic antigens in the Immune Epitope Database (**Figure 6**G and Figure S7A). Additionally, p2 has been detected in multiple cancer types, including glioblastoma, melanoma, and leukemia (Figure S7B). Candidate p10 was completely absent in non-tumor immunopeptidome (Figure S8A), while associated with the MHC of multiple cancer types (Figure S8B). Additionally, we examined both strong and weak ncMAP-HLA interactions predicted by NetMHCpan using DeepImmuno [27]. The DeepImmuno scores estimated the potential for interactions between T-cell receptors (TCRs) and the ncMAP-HLA complexes. Among these, the p2-HLA-B15 complex demonstrated the highest TCR affinity (**Figure 6**H), indicating p2 as the leading immunotherapy candidate identified in this study.

To further demonstrate the evidence of tumor-associated enrichment and recurrent HLA affinities in a clinically relevant background, ELISpot assays were performed on evaluate peptides p2, p6, and p10 using PBMC of HCC patient. A significant increase in IFN-*γ* cytokine release was observed upon exposure to these peptides, further reinforcing their immunogenic potential in HCC patients. Among them, peptide p2 elicited the most pronounced cytokine release response by immune cells, solidifying its position as the leading candidate in this study (**Figure 6**I). These findings underscore the potential of peptide p2 as the most promising immunogenic peptide vaccine candidate, warranting further investigation for therapeutic applications.

### Ribo-Seq and epitranscriptomic data suggest biological mechanisms driving ncMAP overexpression

Elucidating the translational regulatory mechanisms of ncMAPs may yield critical insights for combinatorial treatment strategies. A comparison of TCGA and GTEx data revealed a statistically significant upregulation of PABP and eIF3 subunits in liver cancer tissues compared to normal tissues (**Figure 7**A). Relaxing statistical stringency (p-value *<* 0.05, log2 fold-change *>* 0.5) further revealed significant upregulation of eIF3a, eIF1 and eIF2 subunits, and upstream kinases *EIF2AK1/2* (Figure S9). Notably, p2, p5, p6, and p10 showed a stronger positive correlation with ribosomal subunits relative to other ncORFs (Figure S10). These results suggested association between these ncMAPs and ribosomal subunit expression, meriting deeper investigation of their biological significance. This observation reinforces the involvement of both canonical and non-canonical translation initiation mechanisms in ncMAPs overexpression.

**Figure 7:**
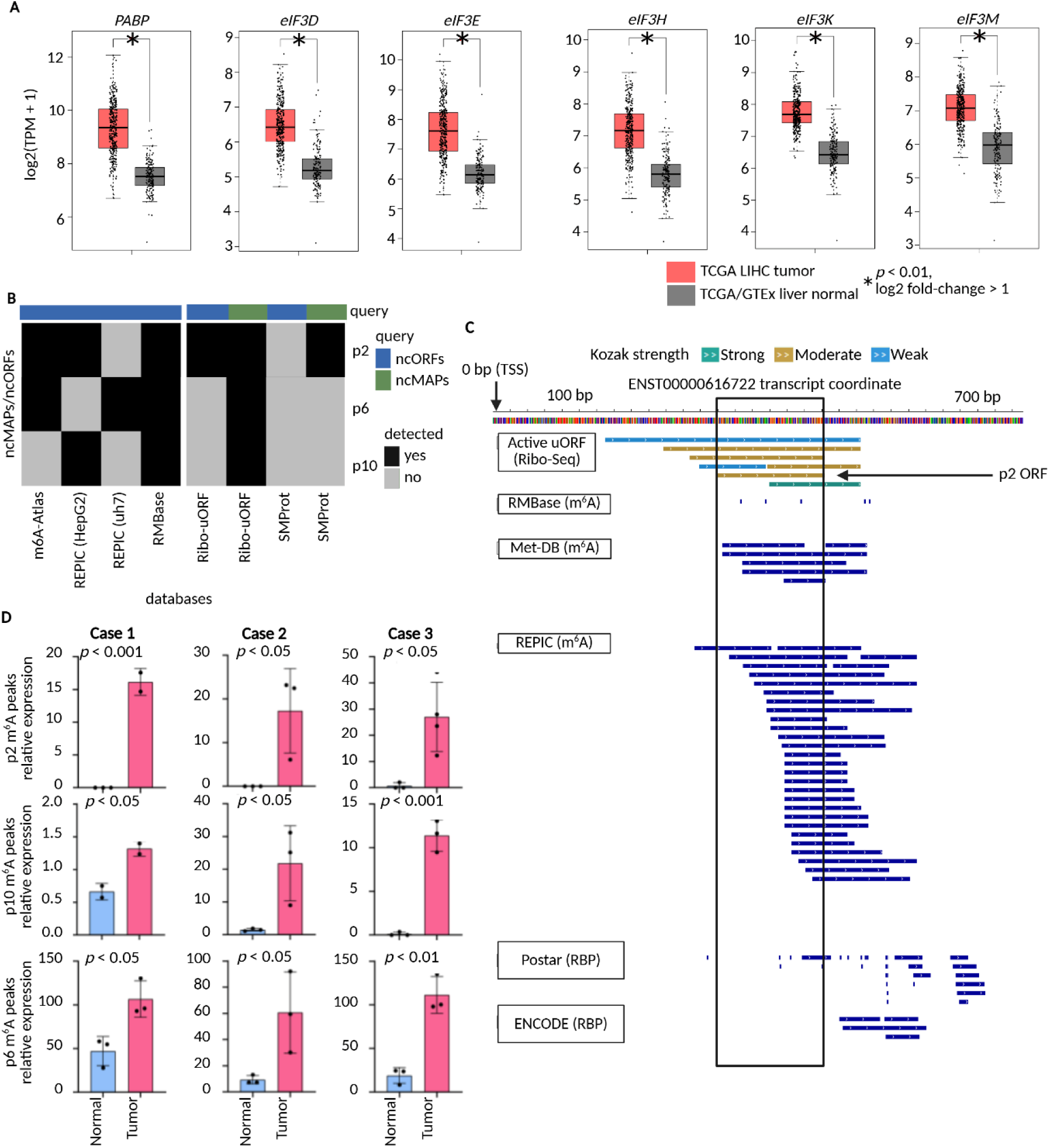
Potential translational regulatory mechanisms of ncMAPs in HCC tissues. **A.** Gene expression of translational regulators in tumor and liver normal tissues from TCGA-LIHC and GTEx, analyzed under stringent statistical criteria (* *p <* 0.01, log2 fold-change *>* 1). **B.** Comparison of peptide and ORF sequences of p2, p6 and p10 against public Ribo-Seq and m^6^A modification datasets. **C.** Integrative analysis of p2 ORF coordinates with Ribo-Seq, m^6^A and RBP annotations from Ribo-uORF. **D.** MeRIP-RT-qPCR quantification of m^6^A methylation levels between HCC tumor tissue and matched adjacent normal tissue of 3 HCC cases. m^6^A. N6-Methyladenosine; RBP, RNA binding protein.; MeRIP-RT-qPCR, Methylated RNA Immunoprecipitation Reverse Transcription Quantitative Polymerase Chain Reaction.

We further investigated the potential regulatory mechanisms underlying ncORF translation initiation using multi-omics data from publicly available databases on HCC cell lines. Our analysis primarily focused on peptides p2, p6, and p10, as they displayed stronger immunogenic potential. The ORFs of p2, p6, and p10 were identified as overlapping with m^6^A peaks across multiple databases. Notably, the REPIC database, which provides cell line-specific annotations for m^6^A peaks, was utilized to query these ncORFs against HCC cell lines Huh7 and HepG2. The results revealed that all three ncORFs exhibited overlap with their respective m^6^A peaks in at least one or both HCC cell lines. Additionally, Ribo-Seq analyses from databases such as RibouORF and SMProt further substantiated the ribosomal recruitment of all three ncMAPs (**Figure 7**B).

To further illustrate the potential regulatory mechanism, the coordinates of the ncORFs of interest were visualized using Ribo-uORF, a database that integrates public datasets of ribosome recruitment, m^6^A modifications and other RNA binding proteins (RBPs). The result identified p2 ORF as an actively translated upstream ORF (uORFs) with moderate Kozak strength, supported by evidence of active translation from Ribo-Seq data in HEK293, glioblastoma, and fibroblast samples. The p2 ORF and other active uORFs of *SERPINB6* coincided with regions containing m^6^A peaks from HCC and other cancer cell lines. Moreover, the p2 ORF overlaps RBP binding sites annotations from Postar and ENCODE. These annotations correspond to binding sites for *DDX3X*, a cytoplasmic scaffold protein known for its interactions with translation initiation factors, ribosomal subunits, and mRNA (**Figure 7**C). Similarly, supporting evidence was observed in regions either overlapping and/or in proximity to ORFs of p6 and p10 (Figure S11A - B). These findings highlight the potential regulatory mechanisms influencing the translation initiation of ncMAPs.

To investigate the potential regulatory role of m^6^A modifications in tumor-associated ncMAPs, we conducted Methylated RNA Immunoprecipitation Reverse Transcription Quantitative Polymerase Chain Reaction (MeRIP-RT-qPCR) quantification of m^6^A peak expression across three matched tumor-normal tissue pairs. The RT-qPCR results showed a significant increase in m^6^A modification for all three ncMAPs across HCC samples relative to their paired normal tissues. Notably, p2 in HCC case 1 and p10 in case 3 exhibited the most pronounced elevation in m^6^A peak expression in tumor tissues (**Figure 7**D). These findings support the functional enrichment of m^6^A modifications in HCC, potentially highlighting the possibility of utilizing m^6^A-modulatings drug to enhance therapeutic efficacy of ncMAP cancer vaccine.

## Discussion

Our study utilized MHC-pulldown mass spectrometry to identify and analyze a collection of ncMAPs obtained from long-read and short-read RNA transcriptomic data. Using a multi-disciplinary approach, we assessed the therapeutic potential of these ncMAPs. Notably, our findings highlighted the 5’-UTR as the primary source of ncMAPs, despite 5’-UTR comprising the smallest portion of the ncORF database in our mass spectrometry data. This pattern is consistent with previous RiboSeq and proteogenomic studies [28, 29, 30]. Peptides derived from 5’-UTRs and other non-coding regions have been shown to play regulatory roles in multiple cellular pathways and have also been presented as neoantigens [31, 32, 33].

We prioritized candidates for further investigation using RNA-level evidence to estimate tumor specificity. While RNA abundance does not directly reflect peptide levels, it is a more reliable metric than proteomics, which is hindered by post-translational modifications, low peptide abundance, and proteasome-mediated degradation. Attempts to quantify our twelve ncORFs across four HCC proteomics datasets detected none. Additionally, ORFs with similar RNA expression levels showed a detection rate below 50% across the same datasets (Figure S12), reinforcing RNA abundance as the most suitable indicator for estimating tumor specificity. Nonetheless, we acknowledge the limitation of using RNA abundance to estimate tumor specificity. In particular, ncORFs derived from frame-shifting events are prone to misquantification because their RNA coverage is indistinguishable from that of the overlapping canonical ORF.

Among peptides that showed evidence of tumor enrichment at RNA levels, p2 has emerged as a strong neoantigen candidate for HCC cancer vaccine development. Further characterization of p2 revealed its strong MHC and TCR affinity, significant HCC enrichment, and ability to trigger CD8^+^ T cell releasing IFN-*γ*. The DeepImmuno prediction suggests p2 is particularly suitable for patients with the HLA-B15 allele, in contrast existing peptide vaccine clinical trials that primarily targeted patients with HLA-A02 and HLA-A24 alleles [34, 35, 36]. Additionally, p6 and p10 demonstrated varying levels of potential as peptide vaccine candidates.

Some of our identified ncMAPs have previously been linked to HCC and immune response. Peptide p2 is derived from the *SERPINB6*, a serine protease inhibitor previously associated with autosomal recessive nonsyndromic sensorineural hearing loss and implicated in cancer-related pathways, including autophagy and apoptosis [37]. *SERPINB6* is also involved in innate immunity, inhibiting cathepsin G (*CatG*)-mediated necrosis and inflammation in neutrophils and monocytes [38]. Peptide p10 originates from *PLOD3*, a key enzyme involved in collagen biosynthesis and thus extracellular matrix remodeling. Targeted inactivation of *PLOD3* has been shown to inhibit liver tumorigenesis [39]. Beyond its functional role, p10 highlights the significance of TE domestication in gene regulatory networks [40, 41]. The *MIR3* element overlaps the 5’ end of the p10 ncORF, consistent with prior research indicating that *MIR* elements can serve as transcriptional promoters and enhancers [42, 43].

While both p2 and p10 showed promising results in terms of immunogenic potential and the absence in normal liver tissues, it is worth noting that IEAtlas data indicated modest p2 presentation in a few brain samples, highlighting the potential challenge of off-target toxicity in certain patients. Although the blood-brain barrier restricts the penetration of immune cells, additional safety measures, such as direct injection or nanoparticle-based localized delivery, should be explored to maximize patient safety [44, 45]. In contrast, p10 was absent from all IEAtlas non-tumor samples, indicating a safer immunogenic profile with minimal risk of autoimmune complications. However, its immunogenic response may be comparatively weaker, as indicated by IEDB, DeepImmuno, and Elispot analyses. It is also worth noting that IEAtlas does not provide definitive proof of the tumor specificity of ncMAPs, as immunopeptidomics mass spectrometry fail to detect low-abundance antigens and can be limited by the efficiency of MHC antibody pulldown. Therefore, IEAtlas should be considered only as supporting evidence, complementing RNA-level analyses to improve the accuracy of assessing of tumor specificity.

PABP and eIF3 subunits showed strong upregulation in TCGA-LIHC samples over GTEx samples. These factors contributed to non-canonical translation initiation by recognizing m^6^A modification [46]. RNA m^6^A modifications were enriched in the p2, p5, p6, and p10 ncORFs, as indicated by public HCC cell line datasets and validated through MeRIP-qPCR results from this study. Additionally, CD8^+^ T cell tumor infiltration has been shown to improve through the activities of the RNA methyltransferase METTL14 and the m^6^A reader proteins YTHDF1 and YTHDF2 [47, 48]. Several m^6^A-modulating anti-cancer drugs, including thapsigargin and tunicamycin, enhance m^6^A writer expression and selectively increase Ribo-Seq coverage for ncORFs [49]. Additionally, Dac51, an inhibitor of the m^6^A eraser FTO, has shown promise in improving the efficacy of anti-PD-L1 blockade in melanoma and non-small cell lung cancer [50]. More recently, oxaliplatin has been reported to elevate m^6^A levels in peritumoral hepatocytes, thereby enhancing anti-PD-1 treatment efficacy against HCC via the YTHDF2-CX3CL1 axis [51]. These findings suggest that the therapeutic potential of peptide vaccine candidates in this study may be enhanced through synergistic modulation of m^6^A modifications.

Our study has identified twelve ncMAPs, with peptides p2 and p10 representing the most promising candidates for peptide-based cancer vaccine. However, as seen in other immunopeptidomic studies, vaccine development is challenged by the limited sensitivity of mass spectrometry, which can lead to misquantification of antigen abundance due to technical constraints such as ionization efficiency, antibody affinity, post-translational modifications, and sample loss. Detection sensitivity can be enhanced using *de novo* search algorithms like PEAKS DB [52], as well as high-resolution mass spectrometry techniques, including data-independent acquisition (DIA) and selected reaction monitoring (SRM). Additionally, expanding the search database to incorporate alternative post-translational modifications and non-ATG start codons [53, 54, 32], along with experimentally validated microproteins [28, 55, 56, 57], and ncORFs predicted through computational methods, such as Splicing Neo Antigen Finder (SNAF) [18], may significantly improve ncMAP identification rates. However, careful database size management is essential, as excessive expansion has been shown to increase false-positive identifications [79]. Besides, this study has a relatively small sample size and the low coverage of mass spectrometry, which likely restricted the number of identifiable peptides and contributed to the absence of ncMAPs mapping directly to splicing junctions, highlighting the need for an expanded mass spectrometry cohort with a greater number of fractionations. Enhancing the detection of ncMAPs from novel splice junction fusions remains a key direction for future research, as these peptides could be promising neoantigen candidates given their sequence divergence from wild-type proteins.

Another limitation of this study is the challenge of clinical translation when using cell line models. These models were selected to enhance ncMAP recovery, as primary tumor tissues often suffer from low tumor purity, which can hinder the detection of tumor-associated antigens. However, patient-derived cell lines do not fully replicate the three-dimensional tumor microenvironment or tumor heterogeneity, limiting their translational relevance. To address this, future research may explore organoid models, which offer a robust platform to overcome the purity limitations of bulk tumor tissues while preserving *in vivo* tumor architecture and cellular heterogeneity. The potential of organoid models was recently demonstrated in a pancreatic cancer study, where MHC-I-associated peptides from ncORFs were successfully identified across multiple patients and recognized by isolated TCRs [58].

## Conclusion

Our study identified twelve ncMAPs, with in-depth downstream analysis of peptides p2, p6 and p10. Among them, p2 demonstrated the strongest immunogenic potential, though its presence in the immunopeptidome of normal brain tissue raises concerns about off-target toxicity. Improving detection sensitivity through expanded databases and advanced computational approaches, alongside the use of organoid models to better capture tumor heterogeneity, could enhance clinical translation. Furthermore, m^6^A-modulating therapies may further refine ncMAP-based immunotherapy, reinforcing their promise as novel cancer vaccine candidates.

## Materials and methods

### Cell culture

We conducted a cell culture experiment using patient-derived HCC cell lines, including HKCI-2 (RRID: CVCL M173), HKCI-4 (RRID: CVCL M175), HKCI-9 (RRID: CVCL SA07), HKCI-10 (RRID: CVCL SA08), HKCI-11 (RRID: CVCL SA10), HKCI-C1 (RRID: CVCL SA00), and HKCI-C2 (RRID: CVCL SA01), as well as a negative control with an immortalized human hepatocyte cell line (MIHA, RRID: CVCL SA11). The culturing conditions were previously described [21, 23].

### Human blood specimens

We obtained peripheral mononuclear cells from healthy donor buffy coat samples via density gradient centrifugation (Cytiva). Afterward, we utilized QIAamp DNA Blood Kits (Qiagen) to isolate DNA for HLA typing with LinkSēq™ HLA-ABCDRDQDP+ 384 kit (One Lambda) on a QuantStudio^TM^ 7 Flex Real-Time PCR System. Human T and dendritic cells (DC) were enriched from PBMCs using a CD8^+^ cell isolation kit and Pan-DC enrichment kit (Miltenyi Biotec), respectively, and subsequently cultured in RPMI-1640 medium (Life Technologies) supplemented with 10% FBS and 100 U/mL of Penicillin-Streptomycin.

### RNA sequencing

In the previous study, PacBio SMRT-seq data and Illumina HiSeq data have been generated for HKCI-2, HKCI-4, HKCI-9, HKCI-11, HKCI-C1, HKCI-C2, and MIHA [21]. In this study, we generated RNA sequencing data for HKCI-10 using the same methodology.

### Affinity purification and LC-MS/MS

MAP-MHC complex was captured by affinity purification in an immunoaffinity column and eluted for liquid chromatography–mass spectrometry identification. See the “Supplementary materials” section for more details.

### Mass spectrometry data analysis

The mass spectrometry data was analyzed using MaxQuant (RRID: SCR 014485, version 2.1.4.0), employing the Andromeda search engine [59]. The search was performed against the human UniProt database (RRID: SCR 002380), and the customized non-reference proteomics database constructed from long-read and short-read RNA-seq data. Briefly, we identify splice junctions from the HCC cell lines by integrating high-throughput Illumina short-read and PacBio long-read sequencing data using the SpliceMap-LSC-IDP algorithm [60]. For a quantitative assessment of alternative splicing, we calculated event inclusion levels, or Percent Splicing Indices (PSIs), using SUPPA (version 2.0.0) [61] with default parameters, filtering for HCC specific splicing events by excluding events with non-zero PSIs in the MIHA cell lines. We further detected splicing events between transposable elements (TE) and exons to expand the ORF candidates in the proteomics search database. TE-exon splicing junctions were identified using STAR chimeric alignment and StringTie2 and transcript assembly. The resulting transcript assembly was then annotated using reference TE and exon coordinates [40]. Additionally, the PacBio reads were analysed with the TEProf2 workflow to identify TE-exon splice variants initiated by TE-derived promoters [41]. Please see the “Supplementary materials” section for more details.

The MaxQuant search parameters were specifically tailored to account for the endogenous digestion process preceding antigen presentation, consistent with similar analyses in the field [62, 63]. This eliminated the need for chemical modifications, such as reduction and alkylation, as well as tryptic digestion during the sample preparation step. Consequently, the search parameters included unspecific digestion and the exclusion of fixed modifications. To accommodate biological post-translational modifications, variable modifications for methionine oxidation and N-terminal acetylation were applied. Moreover, a peptide length range of 8–15 residues were specified, reflecting the typical characteristics of MHC-I-presented peptides. Peptide-spectrum matches (PSMs) underwent statistical filtering, applying a peptide false discovery rate (FDR) threshold of 0.01 and a protein FDR threshold of 1 to account for the limited number of peptides identified per protein following MHC-I enrichment. Several filtering criteria were applied, including removing contaminants and reversed decoy sequences, a minimum MS/MS count of 1, and a maximum posterior error probability (PEP) threshold of 0.01. The hydrophobicity of the filtered peptides was estimated using the R package alakazam [64].

In the re-analysis of datasets PXD023143 and PXD037270, we utilised the same reference and customized proteomics search databases with consistent MaxQuant parameters. The sole exception was the implementation of a more relaxed peptide FDR at 0.05 to enhance the detection sensitivity of MHC-I associated peptides. This adjustment was crucial due to the lower abundance of these peptides in clinical tissue samples, which contain both tumor and non-tumor tissues.

ncMAPs candidates were selected from amino acid sequences exclusively detected in the customized database rather than the UniProt database. The quality of the PSMs of ncMAPs was confirmed by spectrum visualization using the proteomics data viewer PDV [65]. Subsequently, peptides from both UniProt and customized databases were compared against immunopeptidome data available in IEAtlas [19] and HLA Ligand Atlas [66].

### Enzyme-Linked immunospot assay

We evaluated human IFN-*γ* release using the ELISpot Plus Human IFN-*γ* (ALP) kit from Mabtech. See the “Supplementary materials” section for more details.

### Reverse transcription and real-time quantitative polymerase chain-reaction

This study has included Human HCC tumor (T) and adjacent normal liver tissues (AN) from 32 patients. Subjects were eligible for this study if they were qualified for curative HCC surgeries. All surgeries were conducted at Prince of Wales Hospital, The Chinese University of Hong Kong. Before collecting samples, we obtained written informed consent from all patients. The cohort consisted of 27 males and 5 females. Their age ranged from 38 to 78 years old. The patient specimens were processed immediately after surgery and snap-frozen in liquid nitrogen for RNA extraction.

The study utilized the PrimeScript RT Reagent Kit (Perfect Real Time) (Takara, Japan) to reverse transcribe total RNA into cDNA, following the manufacturer’s protocol. The resulting cDNA was either diluted with nuclease-free H_2_O and stored at -80 °C or used directly. Real-time quantitative PCR (RT-qPCR) was performed using the QuantStudio 7 Flex Real-Time PCR system (RRID: SCR 020245). For gene evaluation, the study employed PowerUp SYBR^TM^ Green Master Mix for qPCR (Applied Biosystems) following the manufacturer’s protocol. The expression level of each sample was normalised against 18S rRNA endogenous control for the study was 18S rRNA. Relative expressions were compared across paired T and AN samples to evaluate ncORF overexpression. Primers used in this study were summarized in Table S2.

### Gene expression quantification from RNA-seq cohort

We utilized the gen3-client to download all Genotype-Tissue Expression (RRID: SCR 013042) (GTEx) liver tissue bam files. Meanwhile, all bam files from the Liver Hepatocellular Carcinoma (LIHC) samples of The Cancer Genome Atlas (RRID: SCR 003193) (TCGA) were accessed through the Cancer Genomics Cloud platform [67]. We used featureCounts (RRID: SCR 012919) [68] to quantified gene expression of all bam files (GTEx, TCGA, HKCI short-read and long-read data) against a concatenated GTF file containing RefSeq (RRID: SCR 003496) gene annotation and 12 ncORFs.

### HLA and TCR Affinity Prediction

We filtered the canonical MHC I-associated peptides (cMAPs) identified from mass spectrometry using negative controls. These cMAPs were then compared against patient-specific HLA alleles (4-digit resolution) determined through PCR-based LinkSeq HLA typing, enabling peptide-HLA affinity prediction by the NetMHCpan Server (RRID: SCR 018182) (version 4.1). The HLA binding potential of the query peptide was scored using training dataset from Mass-Spectrometry Eluted Ligands (EL) or quantitative Binding Affinity (BA). The prediction score was ranked against scores obtained from a set of random natural peptides to obtain a percentile rank score (% score). A % score ranging from 2 to 0.5 indicates that the query peptide is a weak binder to the corresponding HLA allele, while a % score below 0.5 suggests that it is a strong binder. A peptide was considered a non-binder to the allele if the % score was above 2 [69]. We then used this information to classify the MAPs-HLA affinity in each HKCI cell line. Specifically, we queried each peptide sequence against all alleles from that cell line and used the lowest % score for the classification. We also employed Gibbs Cluster 2.0 to cluster peptides with consensus binding motifs, which were validated against binding motifs presented in the MHC motif viewer [70, 71, 72].

### Analysis of Translational and epitranscriptomic regulation

To investigate the expression levels of translational regulators, we utilized the Gene Expression Profiling Interactive Analysis 2 (RRID: SCR 018294) (GEPIA2) web server to compare RNA expression data of tumor and non-tumor liver samples from TCGA and GTEx [73]. Moreover, to validate and understand the underlying mechanisms responsible for p1 - p12 translation, the ORF and peptide sequences of p1 - p12 were queried against m^6^A-seq and Ribo-Seq data using the following databases: m^6^A-Atlas v2.0, REPIC, RMBase v2.0, Ribo-uORF, and SMProt [74, 75, 76, 77, 78].

### Methylated RNA immunoprecipitation

m^6^A immunoprecipitation was performed following the SYSY MeRIP-sequencing protocol (https://sysy. com/protocols/protocol-ip-m^6^A-sequencing) with minor modifications. Briefly, equal amount of total RNA from HCC tissue and matched adjacent normal tissue (4 µg) was subjected to fragmentation in a buffer containing 10 mM Tris-HCl (pH 7.0) and 10 mM ZnCl_2_ at 70°C for 6 minutes. Fragmented RNA was purified and mixed with 5 µg anti-m^6^A antibody (Abcam, AB208577, RRID: AB_2916290) in IP buffer (10 mM Tris-HCl pH 7.4, 150 mM NaCl and 0.5% (vol/vol) NP-40) at 4°C for 2 hours followed by incubated with protein-A/G magnetic beads (Sigma-Aldrich 16-663) for additional 3 hours at 4°C. After 4 times of extensive wash steps using IP buffer, the immunoprecipitated m^6^A RNA was separated from protein-A/G-magnetic beads using IP buffer containing 7mM m^6^A disodium salt (MedChemExpress HY-111926,) and 200 Unites of RNasin Plus (Promega N2611) under constant shaking for 1 hour at 4°C. The eluted RNA was purified and subjected for reverse transcription and quantitative real time PCR analysis. Primers used in this study was summarized in **Table S**2.

### Ethics statement

The Joint Chinese University of Hong Kong– Hospital Authority New Territories East Cluster Clinical Research Ethics Committee approved the human sample collection protocol, and informed consent was obtained from each patient. The use of these samples for research was approved by the Joint Chinese University of Hong Kong–New Territories East Cluster Clinical Research Ethics Committee (Ref. No 2020.420).

## Data availability

The mass spectrometry proteomics data have been deposited to the ProteomeXchange Consortium via the PRIDE partner repository with the dataset identifier PXD062193.

## CRediT author statement

**Stephen Li:** Conceptualization, Data curation, Formal analysis, Methodology, Software, Visualization, Writing– original draft, Writing– review & editing. **Yujuan Dong:** Data curation, Investigation, Methodology, Validation, Visualization, Writing– original draft, Writing– review & editing. **Jiaxun Liu:** Investigation. **Shanglin Li:** Validation. **Dandan Pu:** Formal analysis, Software, Visualization, Writing– review & editing. **Yue Guo:** Investigation. **Chun Kwok Wong:** Resources. **Nathalie Wong:** Conceptualization, Data curation, Funding acquisition, Methodology, Project administration, Resources, Supervision, Writing original draft, Writing– review & editing. **Jason Wing Hon Wong:** Conceptualization, Formal analysis, Funding acquisition, Methodology, Project administration, Resources, Supervision, Writing– original draft, Writing– review & editing. All authors have read and approved the final manuscript.

## Acknowledgements

This work was supported by the Hong Kong Research Grants Council Area of Excellence Scheme (Ref. AoE/M-401/20) and an ITC RAISe+ Fund (Ref. RAI/24/1/077A) to Nathalie Wong. The project was also supported by the Research Grants Council (17100920) and the Innovation and Technology Commission, HKSAR, China Mainland-Hong Kong Joint Funding Scheme (MHP/054/21) to Jason W H Wong. LC-MS/MS analysis were performed at LKS Faculty of Medicine, Proteomics and Metabolomics Core Facility, Centre for PanorOmic Sciences, the University of Hong Kong.

## Declaration of generative ai and ai-assisted technologies in the writing process

During the preparation of this work the authors used HKU ChatGPT and Microsoft Copilot to improve the readability and language of the manuscript. After using this tool/service, the authors reviewed and edited the content as needed and take full responsibility for the content of the publication.

