## Supplementary figures for "Discovery and validation of non-canonical antigens for Hepatocellular Carcinoma immunotherapy"

**Supplementary materials**

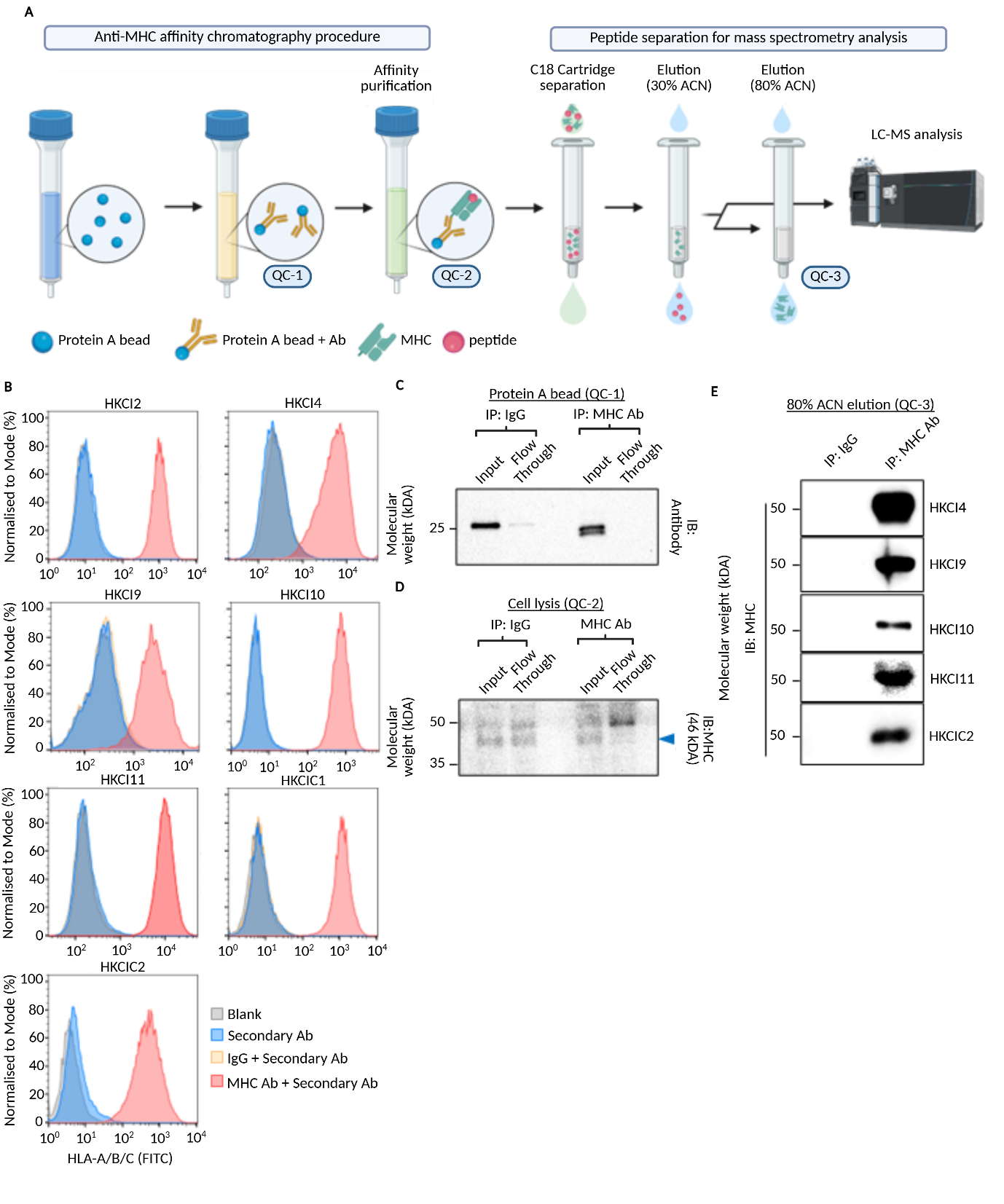
**Figure S1: MHC-A/B/C immunoaffinity purification and antigen enrichment.**

**A.** Graphical summary of MHC-A/B/C immunoaffinity purification and antigen enrichment. Three quality control steps were indicated in the Workflow. Image was created using BioRender software. **B.** HCC cell line surface MHC-class I molecules expression was determined with immunostaining followed by FACS analysis. **C.** The input and flow-through of IgG or MHC-A/B/C antibody after binding with protein A beads was evaluated by western blot analysis. Equal amount of IgG and MHC-A/B/C Ab were used in the immunoaffinity columns. **D.** The MHC complex in the cell lysis before and after binding with immunoaffinity column was determined by western blot analysis. **E.** MHC-A/B/C molecules eluted by 80% ACN was detected by western blot analysis. Representative images were shown. ACN: acetonitrile
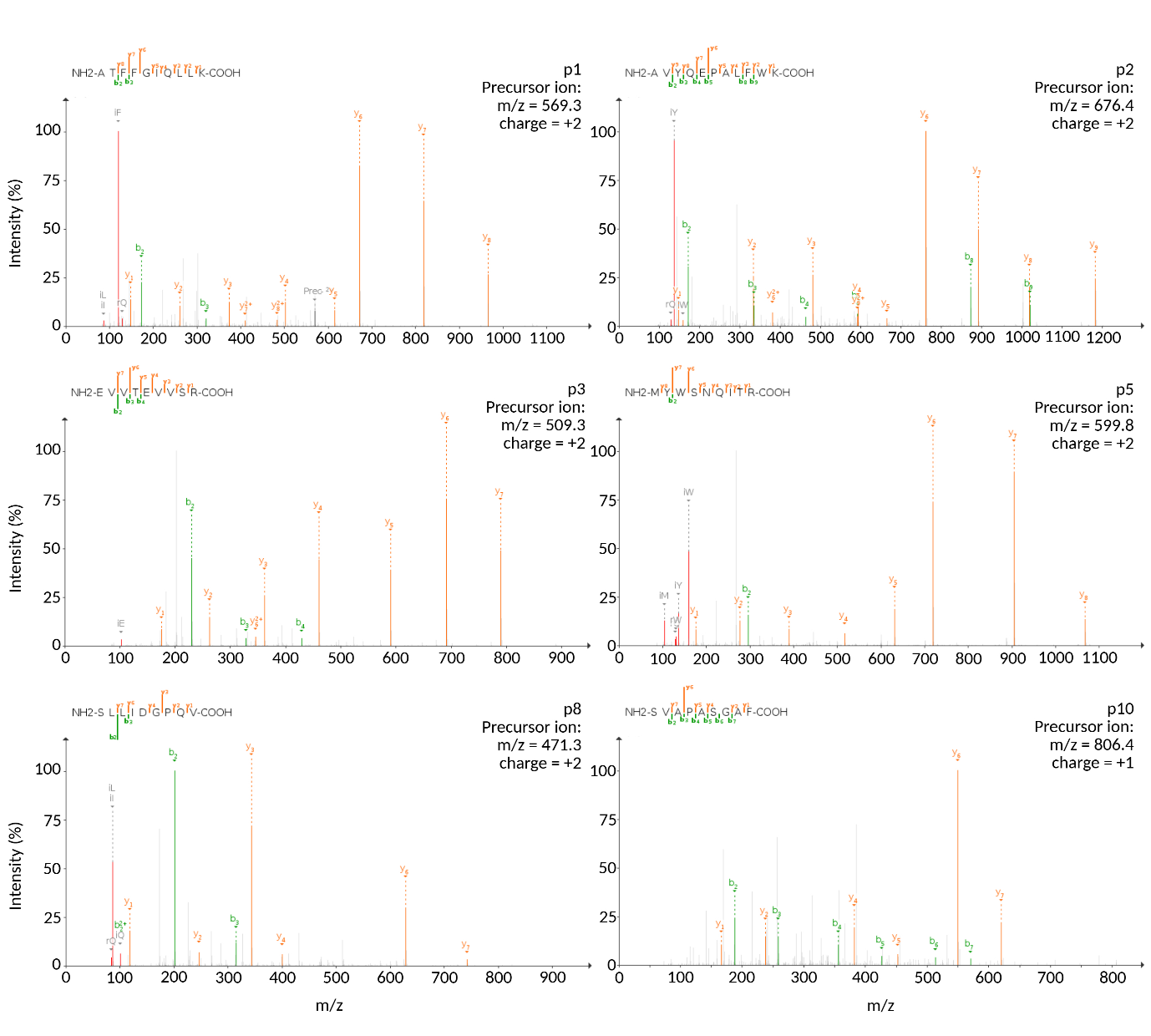
**Figure S2: Representative experimental spectra of ncMAPS and its theoretical spectra.**

All figures were generated using the PDV software to match the mass-to-charge ratio (m/z) between the experimental mass spectra and the theoretical mass spectra derived from the fragmentation of peptide precursor ions. Unmatched peaks are colored grey. Peaks are colored green if they correspond to B ions (fragments containing the N-terminal) and in orange if they correspond to Y ions (fragments containing the C-terminal). The y-axis represents the raw intensity of the mass spectrometry signal, while the x-axis indicates the m/z values. 

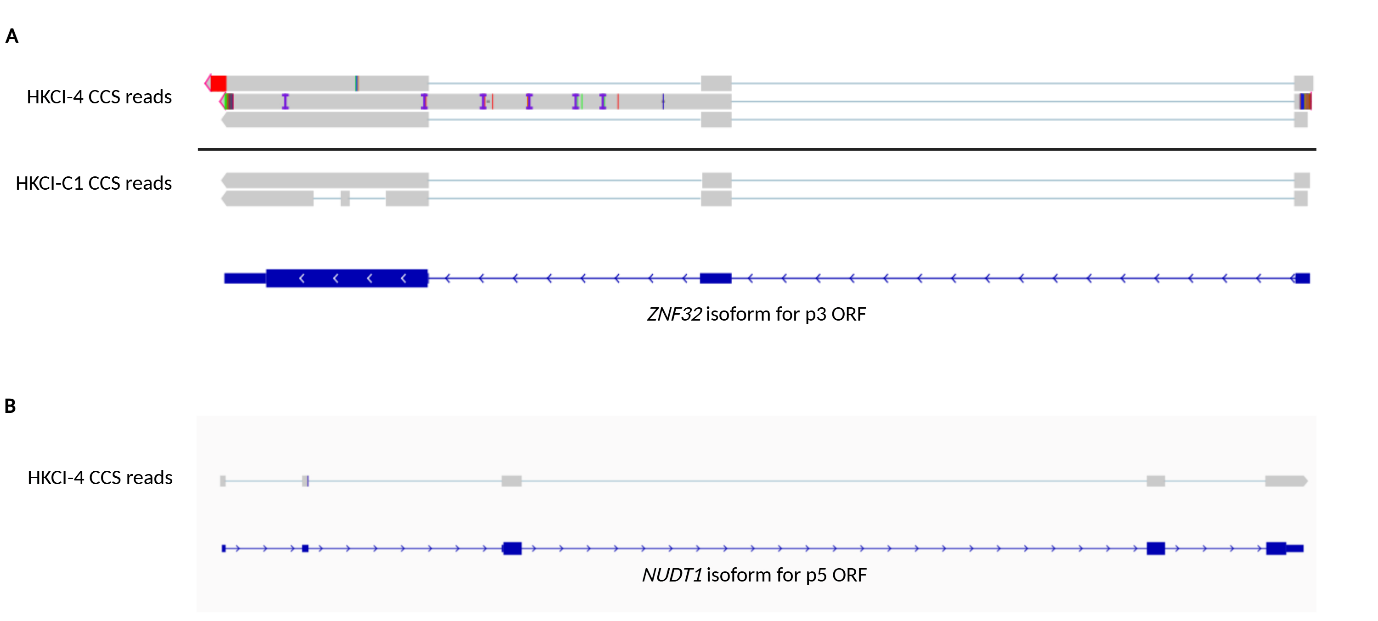
**Figure S3: Visualization of PacBio long-read sequencing data for transcripts associated with p3 and p5.**

**A.** CCS reads from HKCI-4 and HKCI-C1 validating a *ZNF32* isoform, which include the splice junction covered by the p3 ORF. **B.** CCS read from HKCI-4 supporting the *NUDT1* isoform associated with the p5 ORF. CCS, circular consensus sequencing.

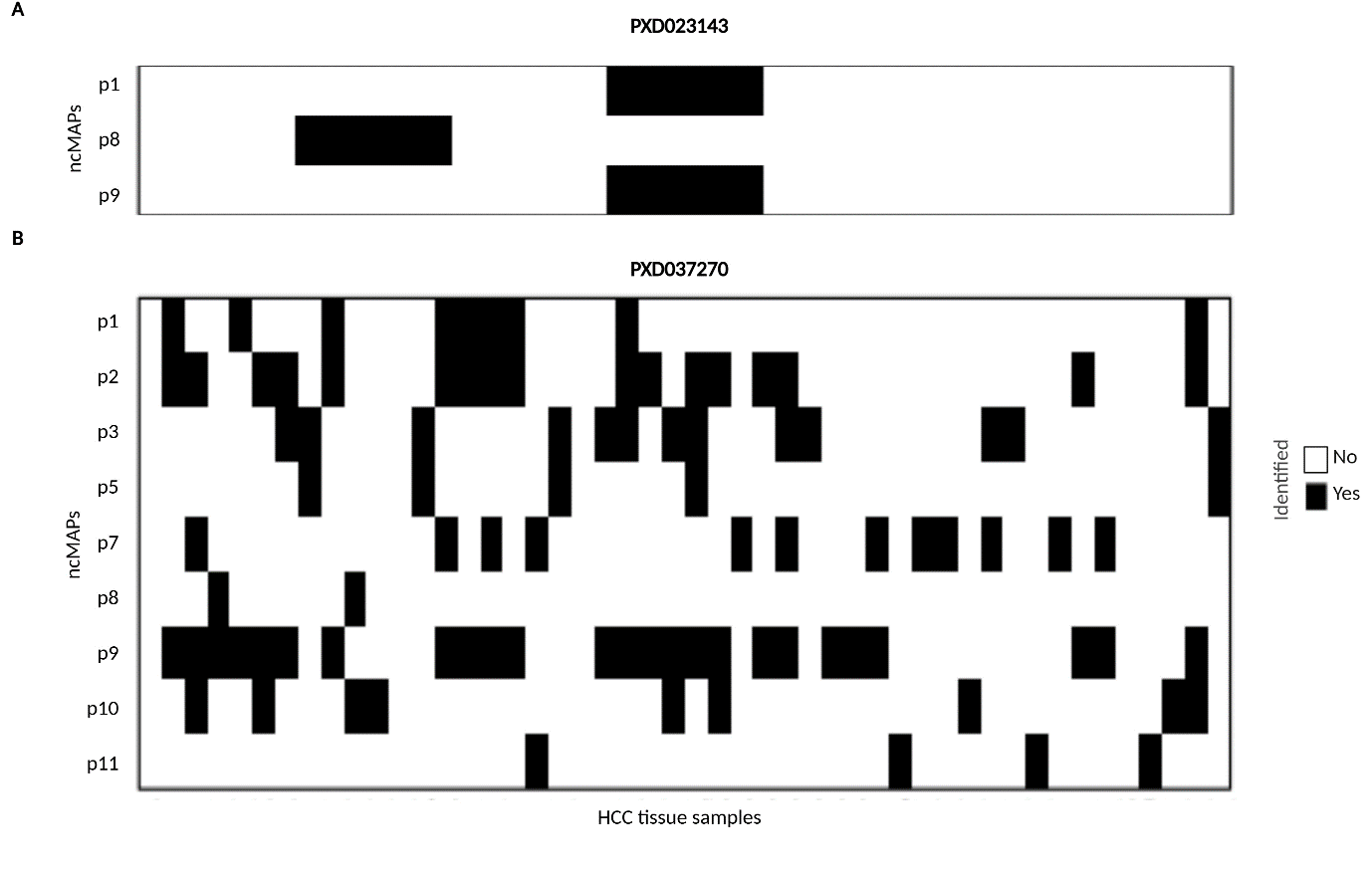
**Figure S4: Identification of ncMAPs in tumor tissues of HCC.**

**A.** The detection of peptides p1, p8, and p9 in dataset PXD023143. **B.** The identification of ncMAPs in dataset PXD037270 across 48 HCC patients.

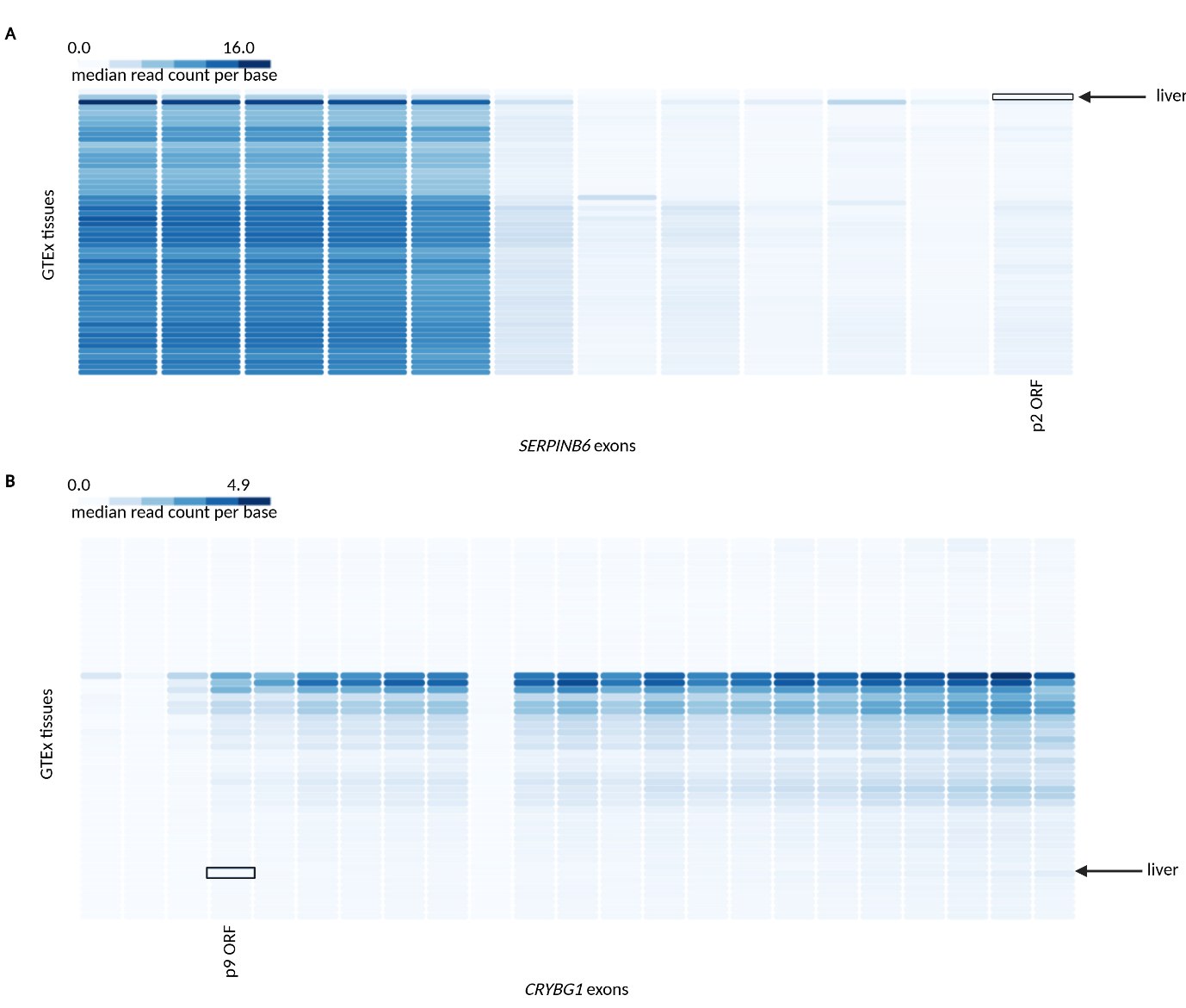

**Figure S5: ncORF expression in human normal tissues at the exon level.**

**A.** Expression of p2 exon in the GTEx dataset. **B.** Expression of p9 exon in the GTEx dataset. The black rectangles highlight the exon of interest in the liver tissue samples. All figures were generated by the GTEx Portal.

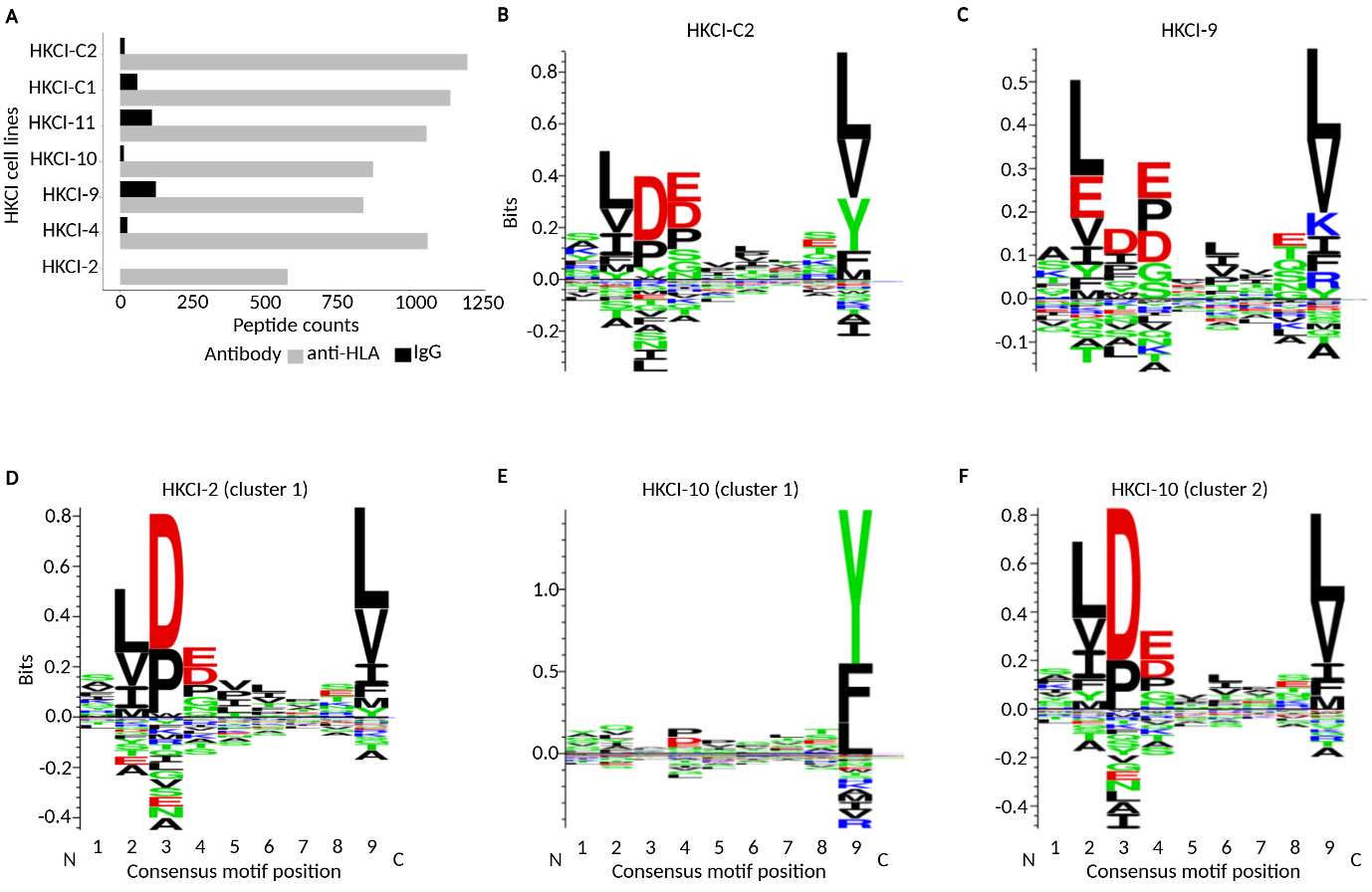
**Figure S6: Summary of consensus motifs of experimentally identified peptides in HKCI cell lines models.**

**A.** The number of peptides captured by anti-HLA and IgG antibodies from each sample for NetMHCpan prediction. **B.** Consensus motif from 1193 peptides detected in HKCI-C2. **C.** Consensus motif from 646 peptides detected in HKCI-9. **D.** Consensus motif from the first cluster of HKCI-2, consisting of 422 peptides. **E.** and **F.** Consensus motif from the two clusters of HKCI-10, consisting of 415 and 558 peptides, respectively. All figures were generated using GibbsCluster 2.0.
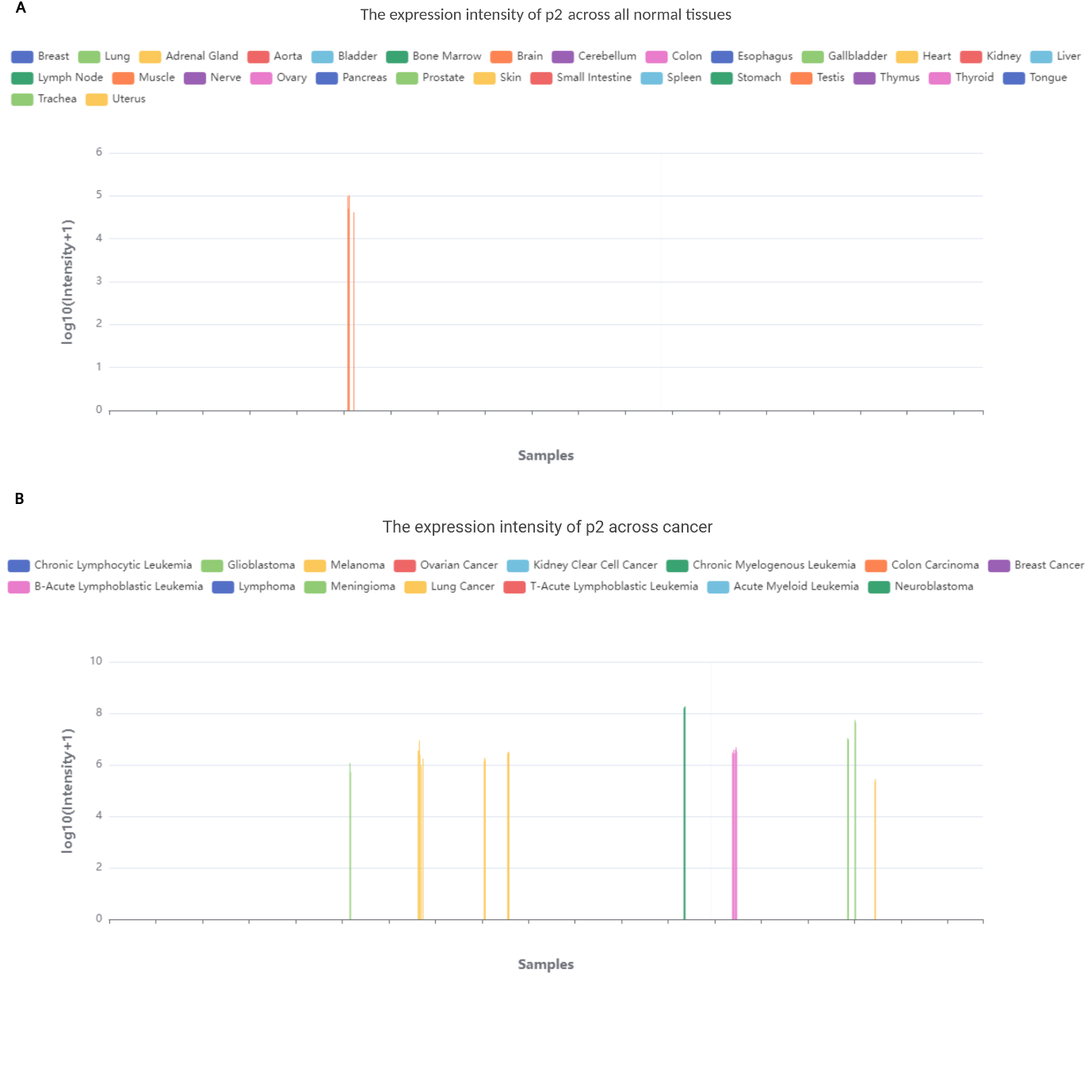
**Figure S7: Pan-Tissue immunopeptidomic profiling of p2.**

**A.** p2 MHC-bound peptide presentation across all normal tissues. **B.** p2 MHC-bound peptide presentation across cancer types. All figures were generated using the IEAtlas.

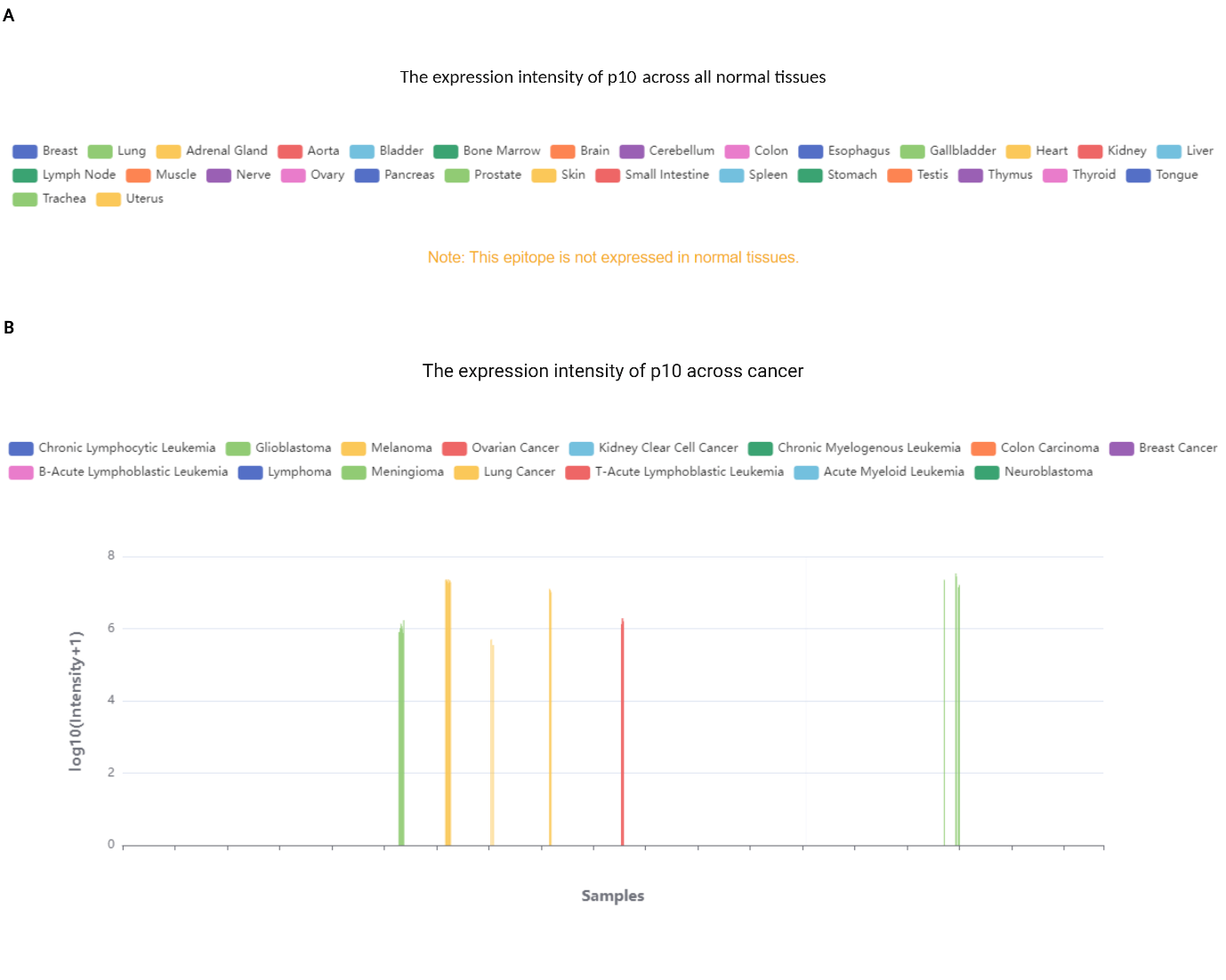
**Figure S8: Pan-Tissue immunopeptidomic profiling of p10.**

**A.** p10 MHC-bound peptide presentation across all normal tissues. **B.** p10 MHC-bound peptide presentation across cancer types. All figures were generated using the IEAtlas.

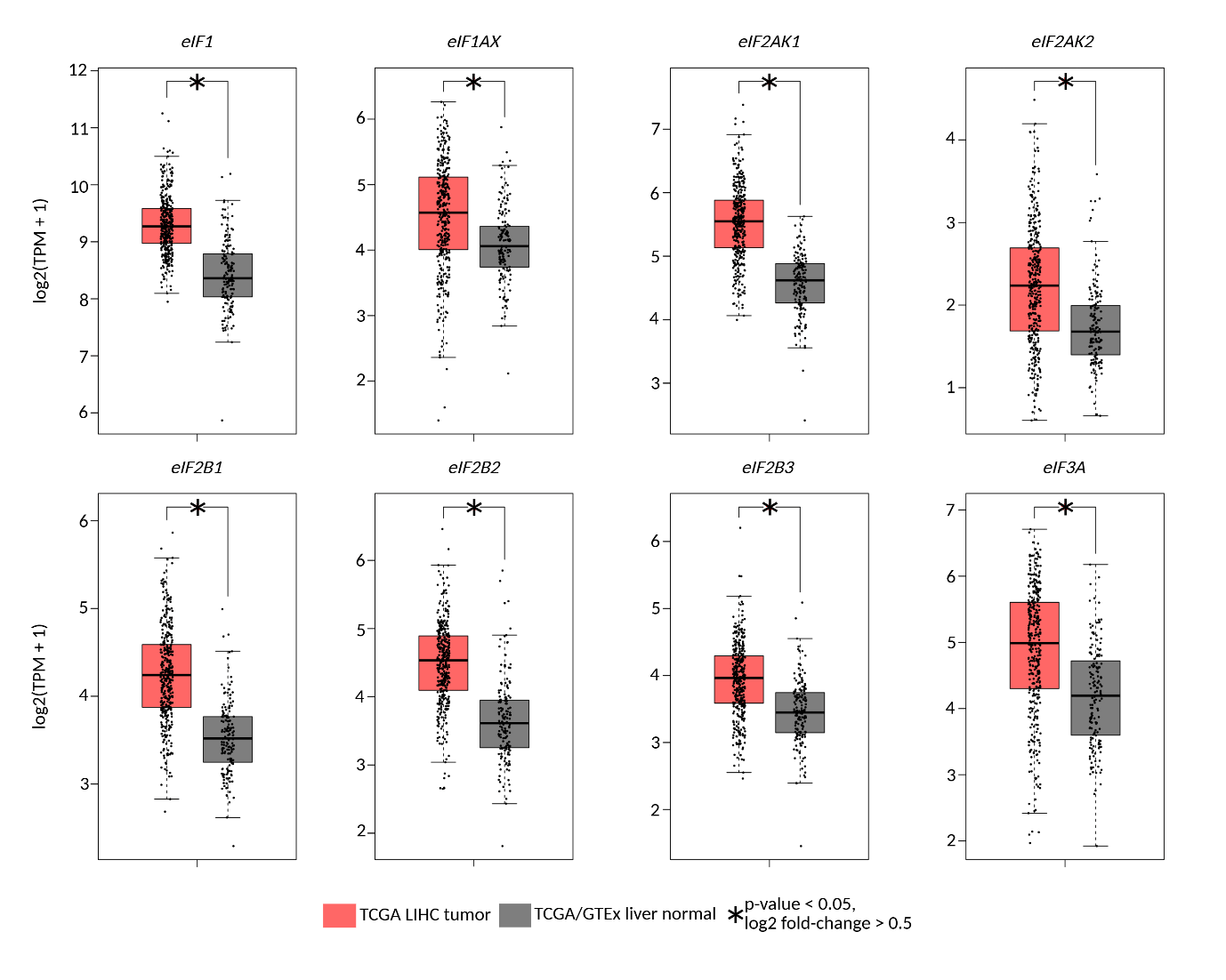

### Figure S9: Translational regulators gene expression in TCGA-LIHC tumor

### and GTEx datasets.

The expression levels were compared with relaxed statistical criteria. **p* *<* 0.05, log2 fold-change *>* 0.5. All figures were generated by GEPIA 2.

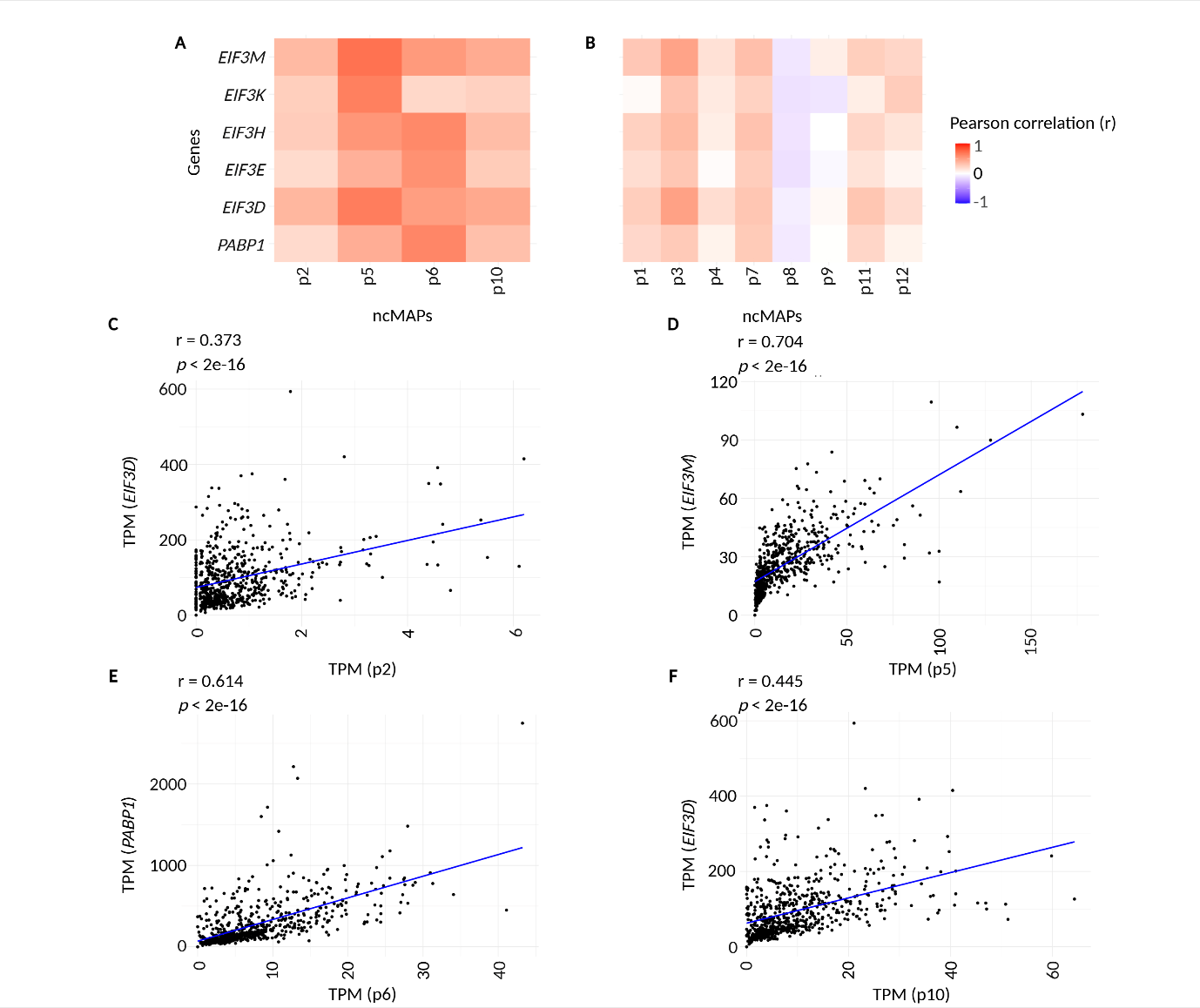
**Figure S10: Correlation analysis of ncMAPs and with the expression of ribosomal subunits.**

A. Pearson correlation between ncMAPs (p2, p5, p6, and p10) with ribosomal subunits.  B. Pearson correlation between other ncMAPs subunits C. to F. Representative correlation scatter plots between selected ncMAPs and ribosomal subunits.

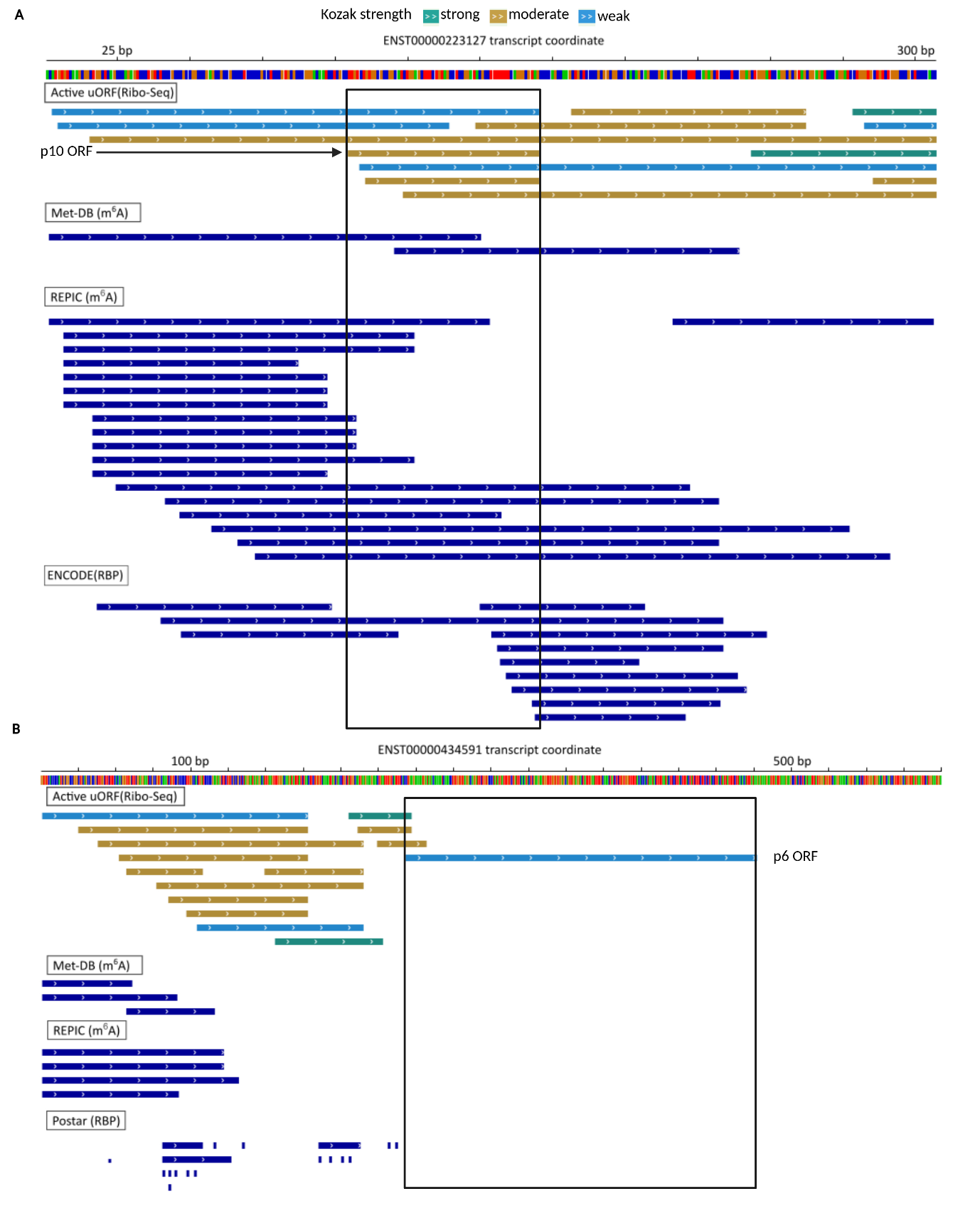
**Figure S11: Visualization of ribosomal uORF regions highlighting m^6^A modification sites and RBP interactions.**

Integrative analysis of A. p10 ORF and B. p6 ORF coordinates with Ribo-Seq, m^6^A and RBP annotations from Ribo-uORF. 

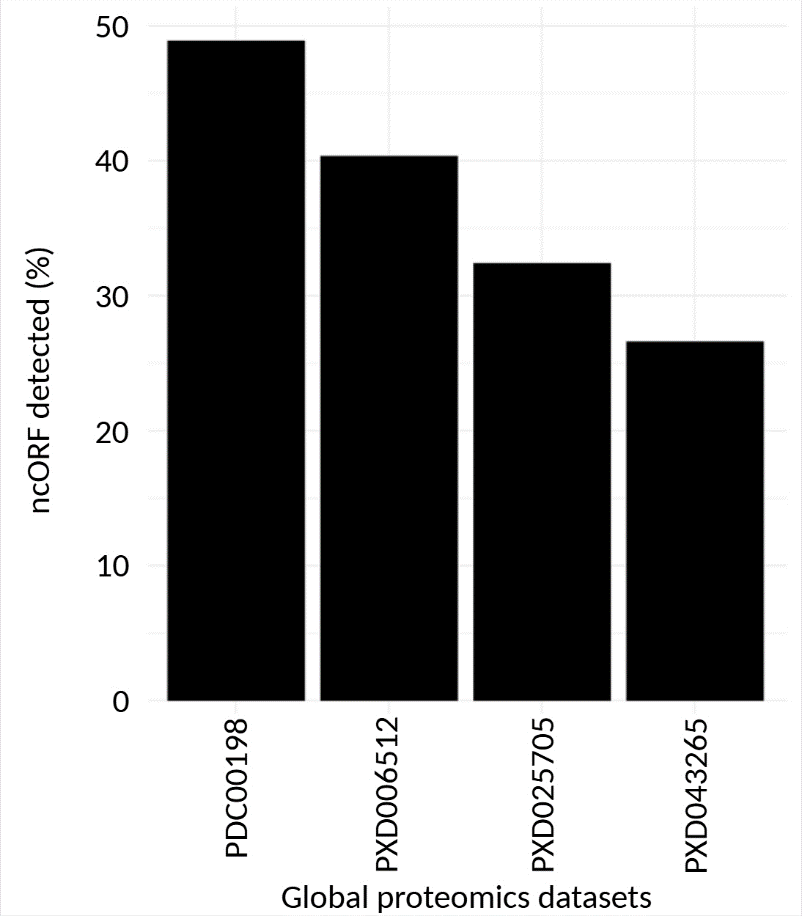

**Figure S12: Mass spectrometry identification of ncORFs across public global proteomics dataset.**

Detection rate of genes with median expression comparable to ncORFs in this study across public HCC proteomics datasets.
