## Supplementary material for "Discovery and validation of non-canonical antigens for Hepatocellular Carcinoma immunotherapy": Table S2

**Table: qPCR primer sets used in this study**

**Name	Sequence**

P2-m6a-Fwd	GACGGTAGCTCGAGACC P2-m6a-Rev	AGACACGTACTCCAGAGC P10-m6a-Fwd	CCAGAGTCCCTGTCTCC

P10-m6a-Rev	GCTACAGAAAGCTCCAGATG P6-m6a-Fwd	GGATTCTGTGCCCTATATGATT

P6-m6a-Rev	ACCCACAAGAGATAGTTTGC

P1-QPCR-Fwd	TGTTGGGAATAAGCTCTGC

P1-QPCR-Rev	ACCTGGAGTTGAGGAATTTG P2-QPCR-Fwd	ATGGCCGTGTATCAGGA

P2-QPCR-Rev	CACCAGCGTGTTTATGGT P3-QPCR-Fwd	ATGAAGAGGTGGTGACTGA P3-QPCR-Rev	CGCTACATTCTGGGCATATC P4-QPCR-Fwd	AGCGGCCAAGCTCAT

P4-QPCR-Rev	CCGTCCCAGTAGTCTTCAG P5-QPCR-Fwd	GTACTGGAGCAATCAGATCAC P5-QPCR-Rev	ACCAGCACCAGGGTATAG

P6-QPCR-Fwd	GAGCAGTTGGAGGACTATTTG

P6-QPCR-Rev	GGTAGAGAAGAAAGATAGTAAAGTAAGG P7-QPCR-Fwd	ATGGCTGAGCGTGAAGA

P7-QPCR-Rev	TCAGAGAGGCCGGAGA

P8-QPCR-Fwd	GCTTGTTAATTGATGGACCAC P8-QPCR-Rev	CCCATGCCATTTACTTACTCT P9-QPCR-Fwd	TGAGCACAATCCATTTAGCC P9-QPCR-Rev	AGTGTGTCTTGGGCTACT P10-QPCR-Fwd	CCAGAGTCCCTGTCTCC

P10-QPCR-Rev	GCTACAGAAAGCTCCAGATG P11-QPCR-Fwd	ACAGCCATGAGTCTCACA P11-QPCR-Rev	TAGGGCAAAGGTCTCAGAA P12-QPCR-Fwd	CACGCACGCCAAGAT

P12-QPCR-Rev	TTAGACTGTGGTTACAAGGTG

**Table Note:** This table lists the **qPCR primer sets** used in this study. Primer names containing "**m6A**" indicate primers designed for **MeRIP-RT-qPCR,** used for analyzing **m^6^A-modified RNA enrichment.** Those labeled with "**QPCR**" correspond to primers used for **RT-qPCR**, specifically for **quantifying ncORFs.** All primer sequences are detailed to ensure clarity in experimental setup. MeRIP-RT-qPCR, Methylated RNA Immunoprecipitation Reverse Transcription Quantitative Polymerase Chain Reaction; **ncORFs , non-canonical open reading frames;** RT-qPCR, Quantitative reverse transcription polymerase chain reaction.
