## Supplementary material for "Discovery and validation of non-canonical antigens for Hepatocellular Carcinoma immunotherapy": File S1

**Affinity purification of HLA class I molecules and enrichment of HLA-bound peptides.**

To generate the HLA class I immunoaffinity column, Ultra-LEAF^TM^ Purified anti-human HLA-A/B/C anti body (RRID: AB 314871) (clone W6/32, 311441, BioLegend, San Diego, CA) was incubated with Protein A Sepharose^TM^ 4B beads (101041, Thermo Fisher Scientific) for 12 hours at room temperature. Ultra-LEAFTM Purified Mouse IgG2a κ isotype antibody (RRID: AB 2927399) (clone MOPC-173, 400264, BioLegend) was used as a negative control. Following antibody crosslinking to Protein A Sepharose^TM^ 4B beads in fresh 40 mM dimethyl pimelimidate (DMP) (21667, Thermo Fisher Scientific) in Sodium Borate Buffer (0.2 M H_3_BO_3_, 0.05 M NaOH, pH 8.5) for 1 hour at room temperature, the reaction was terminated using ice-cold Tris buffer (0.2 M Tris, pH 8.0). Unbound antibody was eliminated from the column by extensive washing using a stripping buffer (0.1 M citrate, pH 3.0).

To improve the detection rate of presented epitopes by mass spectrometry, 1.9 × 10^9^ cells were used in each affinity purification. Cell lysate supernatant was generated from freshly prepared cells by homogenizing in lysis buffer (1% NP-40, 150 mM NaCl, proteinase inhibitor (A32963, Thermo Fisher Scientific), 50 mM Tris, pH 8.0) followed by ultracentrifuge (100,000 g for 75 minutes at 4°C). After pre-clearing with Protein A Sepharos^TM^ 4B beads, the lysate was incubated with an immunoaffinity column for 16 hours at 4 °C. Non-specifically bound materials, detergent, and salts was washed from the immunoaffinity column by firstly with 10× column volume of wash buffer 1 (0.005% NP-40, 150 mM NaCl, 5 mM EDTA, 50 mM Tris, pH 8.0), followed by 10 × column volume of wash buffer 2 (150 mM NaCl, 50 mM Tris, pH 8.0) and wash buffer 3 (450 mM NaCl, 50 mM Tris, pH 8.0), and finally with 10 × column volume of wash buffer 4 (50 mM Tris, pH 8.0). HLA-antigen complex was eluted with 0.2% trifluoracetic acid (85183, Thermo Fisher Scientific) from the affinity column into a Protein LoBind 1.5mL microcentrifuge tube (90410, Thermo Fisher Scientific). Antigen peptides were further separated from HLA molecules on a Sep-Pak tC18 Plus Light Cartridge (WAT036805, Waters, Milford, MA) with 30% acetonitrile (10501014, Thermo Fisher Scientific) in 0.1% trifluoroacetic acid and subsequently lyophilized with CentriVap vacuum concentrator (7983015, Lanconco, Kansas City, MO) for mass spectrometry analysis. The remaining HLA molecules were eluted from the Sep-Pak tC18 cartridges with 80% acetonitrile in 0.1% trifluoroacetic acid or western blot detection. Chemicals and solvents used in the wash and elution steps were in LC-MS grade (Figure S1A).

**Liquid chromatography-mass spectrometry (LC-MS/MS) analysis of MHC class I antigen peptides.**

The Dionex Ultimate3000 nanoRSLC system (RRID: SCR 019840), coupled with a Thermo Fisher Orbitrap Fusion Tribrid Lumos mass spectrometer (RRID: SCR 020573), was used to analyze eluted antigen peptides. Peptides were separated on a commercial C18 column (75µm i.d. × 50 cm length × 2µm particle size) coupled to a NanoTrap column (75µm i.d. × 2 cm length × 3µm particle size) (Thermo Fisher). Separation was attained using a linear gradient of increasing buffer B (80% acetonitrile and 0.1% formic acid) and declining buffer (0.1% formic acid) at 300 nL/minute. Buffer B was increased to 30% in ca. 30 minutes and ramped to 44% in the next 15 minutes, followed by a quick ramp to 95%, where it was held for 3 minutes before a quick ramp back to 3%. The column was held and re-equilibrated at 3%. The mass spectrometer was operated in positive polarity mode with a capillary temperature of 300 °C. Full MS survey scan resolution was set to 120,000 with an automatic gain control (AGC) target value of 5 × 10^4^, maximum ion injection time (IT) of 50 ms, and a scan range of 350 – 1500 m/z. Charge states from 1^+^ to 4^+^ were acquired. The dependent scan on single charge state per precursor only setting was also enabled. A data-dependent top speed method with a time interval of 2 seconds between every survey scan was operated, during which higher energy collisional dissociation (HCD) was used. MS/MS spectra were obtained at 60,000 MS2 resolution with a normalized AGC target of 200% and maximum ion injection time of 100 ms, 1.6 m/z isolation width, and normalized collisional energy of 30. Preceding precursor ions targeted for HCD were dynamically excluded for 10 seconds. All solvents were equivalent to or higher than LC-MS grade.

**Proteomics database construction from transcriptomic data.**

Long-read supported splice-variants were identified previously [1]. Tumor-specific variants were extracted by removing isoforms expressed in MIHA. Additionally, fusion transcripts between TE and exons were detected from short-read RNA-Seq data using custom scripts [2]. Moreover, long-read RNA-seq data was processed by Iso-Seq clustering, LSC short-read correction, and Minimap2 alignment [3; 4; 5]. The aligned reads were analyzed by TEProf2 workflow to detect TE-derived alternative transcripts [6].

From the alternative transcripts identified from splice-variants and TE-exon fusions, the ORFik R package detected open reading frames in alternative transcripts ORFs that commence with ATG and are at least 25 amino acids long [7]. The resulting ncORFs, along with the reference UniProt database (RRID: SCR 002380), were combined to form a comprehensive database for subsequent proteomic searches.

**Enzyme-Linked Immunospot assay.**

Briefly, dendritic cells (DCs) were pulsed with a 10 µg/mL peptide concentration at a cell density of 10^5^ cells/mL for 4 hours at 37°C. Following this, the DCs were mixed with autologous CD8^+^ T cells at a DC:T cell ratio of 1:10. The cell mixture was then aliquoted into an Enzyme-Linked Immunospot (ELISpot) plate (3420-4APT-10, Mabtech, Stockholm, Sweden) at 100 µL per well and incubated for 24 hours. The secreted IFN-*γ* signal was detected and quantified according to the manufacturer’s instructions. All peptides used in this study were synthesized by GenScript Biotech (Piscataway, NJ) and had a purity greater than 98%. The trifluoroacetic acid residue was less than 1%, and the endotoxin level was at most 0.01 EU/µg. The anti-human CD3 mAb (CD3-2), a component of the kit ELISpot Plus: Human IFN-*γ* (ALP), was used as a positive control. Another positive control in this study was NY-ESO-1, a cancer-testis antigen extensively studied in immune-based clinical trials. To eliminate false-positive immune signals from potential contaminants during peptide synthesis, we synthesized a peptide derived from human Actin within the amino acid region 1-9 (Actin_1-9_) as a negative control. The amino acid sequences of NY-ESO-1 and Actin_1-9_ are SLLMWITQV and MCDEDETTA, respectively. Prediction with the NetMHCpan EL 4.1 program revealed that the Actin_1-9_ peptide had a relatively low binding affinity with all HLA-I molecules and a low probability of eliciting an immune response (HLA-I affinity score: Actin_1-9_ = 0.006, NY-ESO-1 = 0.943). The ELISpot analysis presented the IFN-*γ* count as the mean ± standard deviation (SD). Statistical analysis was performed using an unpaired Student’s t-test, which defined the statistical significance as *p* below 0.05. We conducted all statistical analyses using GraphPad Prism version 10 (RRID: SCR 002798).
